# Estimating Time-Inhomogeneous Transition Probabilities from District Level Daily COVID-19 Transmission Data in Sierra Leone

**DOI:** 10.64898/2026.07.28.26359161

**Authors:** Abass Jah, Oscar Ngesa, Charity Wamwea, Antony Ngunyi

## Abstract

Compartmental epidemic models conventionally treat the probability of moving between disease states as fixed over time, an assumption that sits uneasily with the reality of a pandemic in which lockdowns, mask mandates, vaccination roll-out, and the arrival of new variants continually reshape transmission. This paper develops a time-inhomogeneous Markov chain framework for the Susceptible–Exposed–Infectious–Removed (SEIR) process, in which each transition probability *p_ab_*(*t*) is allowed to vary with calendar time while respecting the structural zeros implied by the SEIR compartmental flow. We derive the constrained maximum-likelihood estimator of *p_ab_*(*t*) under these structural constraints, establish its finite sample efficiency, asymptotic normality, and Wilson score confidence intervals, and construct a likelihood ratio test of the null hypothesis that a compartment’s exit probability is constant over time. We further propose a stochastic machine learning hybrid extension in which the raw, kernel smoothed transition probabilities are regressed on policy and mobility covariates using both a logistic generalized linear model and a random forest, allowing the framework to attribute time-inhomogeneity to observable interventions. The methodology is applied to a compiled daily, district level COVID-19 surveillance panel for Sierra Leone spanning March 2020 to December 2023 (16 districts, 1,401 days). The likelihood ratio test rejects time-homogeneity of the exposed to infectious transition in 15 of 16 districts and of the infectious-to-removed transition in 8 of 16 districts (*α* = 0.05), and the covariate augmented logistic model achieves an out of sample Brier score roughly 76 times smaller than a time-homogeneous pooled baseline, with healthcare capacity and the time trend emerging as the most influential predictors in the random-forest component. These results provide statistical evidence that time-inhomogeneous, covariate informed Markov models offer a materially better description of district-level COVID-19 transmission in Sierra Leone than classical time-homogeneous compartmental models, with implications for sub-national outbreak monitoring in resource constrained settings.

## 1 Introduction

Compartmental models of infectious disease transmission trace their lineage to the deterministic Susceptible–Infectious–Removed (SIR) framework of Kermack and McKendrick (1927), which partitions a population into mutually exclusive epidemiological states and describes the flow between them through a system of differential (or, in discrete time, difference) equations. Extending the framework by an Exposed compartment yields the SEIR model, which more realistically captures the latent period between infection and infectiousness that is characteristic of SARS-CoV-2. In its most common form, the compartmental model whether written as a system of ordinary differential equations or as a discrete-time Markov chain assumes that the probability of transitioning from one compartment to the next is constant over the course of the epidemic. This assumption of time-homogeneity is a mathematical convenience rather than an epidemiological reality: transmission and progression rates shift as governments impose or relax lockdowns and mask mandates, as population mobility and healthcare capacity fluctuate, as vaccination coverage rises, and as more transmissible or more severe variants of concern displace earlier lineages.

A time-inhomogeneous Markov chain, in which the transition matrix *P* (*t*) is permitted to vary with time *t*, offers a natural and mathematically tractable relaxation of this assumption while preserving the interpretability of a discrete state compartmental model. The central statistical challenge is that, once *p*_*ab*_(*t*) is allowed to change on every time step, there are in principle as many parameters as there are days in the study period, so that estimation without some form of regularization (temporal smoothing) or without imposing known epidemiological structure (structural zeros forbidding biologically impossible transitions, such as a recovered individual reverting to susceptible) is not statistically well posed. This paper addresses that challenge directly: we impose the SEIR structural zero pattern on the transition matrix, derive the constrained maximum likelihood estimator (MLE) of the resulting one step transition probabilities, and develop a full asymptotic inference toolkit efficiency, asymptotic normality, confidence intervals, and a formal likelihood ratio test of time-homogeneity around that estimator. We then extend the purely stochastic model into a stochastic machine learning hybrid by regressing the estimated transition probabilities on policy and mobility covariates using both a parametric logistic model and a non-parametric random forest, in the spirit of recent hybrid compartmental/machine learning approaches to COVID-19 forecasting (Watson et al., 2021; Galasso et al., 2022; Baccega et al., 2024; Zhang et al., 2023).

Sierra Leone offers a particularly instructive setting for this methodology. As a low resource West African country with a comparatively low reported COVID-19 caseload relative to its population (Liu et al., 2022), successive waves of the pandemic were shaped by imported cases, evolving non-pharmaceutical interventions, and the sequential introduction of variants of concern (Lin et al., 2022), all of which plausibly induced substantial time variation in district level transmission dynamics. Because national aggregates can mask heterogeneous, district specific epidemic trajectories, we work directly with a daily, district level surveillance panel (16 districts, March 2020–December 2023) and estimate district specific, time varying transition probabilities rather than a single national curve.

The contributions of this paper are threefold. First, we present a complete constrained likelihood and asymptotic inference framework for time-inhomogeneous, structurally constrained SEIR Markov chains, including a novel bandwidth selection diagnostic based on a Pearson goodness of fit statistic for the kernel smoothed estimator. Second, we introduce a hybrid stochastic machine learning extension that regresses the estimated transition probabilities on interpretable policy and mobility covariates, bridging the classical Markov chain literature with the modern machine learning for epidemiology literature. Third, we provide a full empirical application to district level Sierra Leone COVID-19 data, including a reproducible data reconstruction pipeline that converts routinely reported daily case, recovery, and death counts into SEIR compartment occupancy and flow counts suitable for the proposed estimator.

The remainder of the paper is organized as follows. Section 2 reviews related work on Markov chain epidemic models, multistate statistical models, and hybrid machine learning approaches to epidemic forecasting. Section 3 develops the full mathematical methodology. Section 4 describes the data and the experimental design implemented in R. Section 5 reports the results. Section 6 discusses the epidemiological and methodological implications, and Section 7 concludes.

## 2 Related Work

### Compartmental and Markov chain epidemic models

The deterministic compartmental model of Kermack and McKendrick (1927) remains the conceptual starting point for almost all subsequent work in mathematical epidemiology, and its stochastic, discrete state analogue a continuous or discrete-time Markov chain on a finite set of epidemiological states has been used extensively during the COVID-19 pandemic. Bagyan and Richards (2022) present a continuous-time Markov chain model for COVID-19 spread pedagogically motivated by cruise ship out-breaks; Oka et al. (2020) develop a multivariate discrete-time Markov model of COVID-19 propagation across Chinese provinces that explicitly incorporates human mobility covariates, closely paralleling the covariate augmented extension we propose in Section 3.6. These studies, like the vast majority of Markov chain epidemic models, treat the transition mechanism as time-homogeneous (or homogeneous within short estimation windows), which is the assumption we relax.

### Multistate statistical models and non-parametric transition estimation

Outside epidemiology, the estimation of time varying transition probabilities in a multistate stochastic process has a long history in biostatistics. Aalen and Johansen (1978) derive the non-parametric (Aalen Johansen) estimator of the transition matrix for a non-homogeneous Markov chain under right censoring, which is the direct ancestor of the constrained, row wise maximum likelihood estimator we derive for the SEIR chain. Cook and Lawless (2018) provide a comprehensive modern treatment of multistate models for life history data, including structural zero (forbidden transition) constraints of exactly the kind imposed by the SEIR flow *S → E → I → R*. Our contribution relative to this literature is to specialize the general multistate machinery to the SEIR structural zero pattern, to show that the resulting row wise multinomial likelihood collapses to a binomial likelihood for each non-absorbing compartment, and to build the full efficiency/normality/testing toolkit around this simplification.

### Hybrid stochastic machine learning approaches

A growing literature couples classical compartmental or Markov chain models with machine learning components to combine mechanistic interpretability with data adaptive flexibility. Watson et al. (2021) fuse a Bayesian case velocity model with a random forest (Breiman, 2001) to forecast COVID-19 deaths across U.S. states; Galasso et al. (2022) use a random forest driven by reproduction number estimates and demographic covariates for regional case forecasting; Baccega et al. (2024) integrate compartmental models, machine learning, and variant information within a single forecasting framework; and Zhang et al. (2023) combine an autoregressive component with an LSTM network to retain interpretability while improving predictive accuracy. Our hybrid model differs from this literature in scope: rather than forecasting case counts directly, we use machine learning to model the individual transition probabilities of a mechanistic time-inhomogeneous Markov chain, so that the resulting hybrid retains the SEIR compartmental structure (and its associated inferential guarantees) while allowing the exit probabilities themselves to depend flexibly on policy and mobility covariates.

### COVID-19 epidemiology in Sierra Leone

Descriptive epidemiological work on COVID-19 in Sierra Leone documents a comparatively low but wave structured epidemic curve, imported case driven transmission, and the sequential dominance of different SARS-CoV-2 lineages across successive waves (Liu et al., 2022; Lin et al., 2022). These studies motivate, but do not themselves formally test, the hypothesis of time varying transmission dynamics that we address directly through the likelihood ratio test developed in Section 3.5.

## 3 Methodology

### 3.1 Time-Inhomogeneous Markov Chain Formulation

Let (Ω, *F*, Pr) be a probability space and let 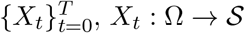, be a discrete-time stochastic process on the finite SEIR state space

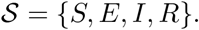

We assume {*X*_*t*_}satisfies the time-inhomogeneous Markov property

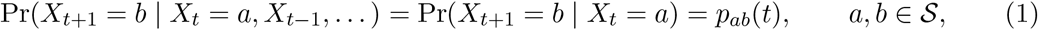

so that the transition mechanism *p*_*ab*_(*t*) is permitted to change with calendar time *t*, reflecting non-pharmaceutical interventions, vaccination roll out, mobility changes, and variant turnover. The time varying transition matrix *P* (*t*) = (*p*_*ab*_(*t*))_*a,b∈S*_ is row-stochastic, ∑_*b*_*p*_*ab*_(*t*) = 1, and the marginal state distribution evolves as *π*_*t*+1_ = *π*_*t*_*P* (*t*).

### 3.2 Structural Zeros and the Constrained Likelihood

The SEIR compartmental flow *S → E → I → R* imposes epidemiologically necessary structural zeros: a susceptible individual cannot move directly to *I* or *R*; an exposed individual cannot revert to *S* or advance directly to *R*; an infectious individual cannot revert to *S* or *E*; and *R* is absorbing. Formally, for all *t*,

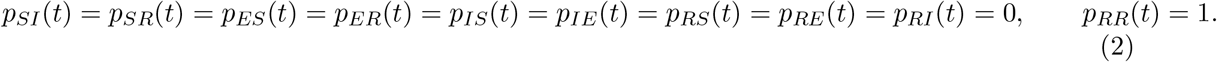

Defining the admissible next state sets *A*(*S*) = {*S, E*}, *A* (*E*) = *{E, I}, A* (*I*) = *{I, R}, A* (*R*) = *{R}*, the transition matrix collapses to

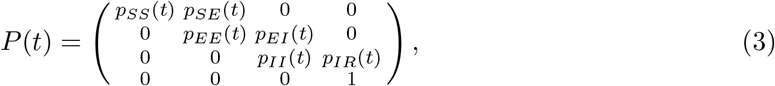

which is row stochastic if and only if *p*_*SS*_(*t*)+*p*_*SE*_(*t*) = 1, *p*_*EE*_(*t*)+*p*_*EI*_(*t*) = 1, *p*_*II*_(*t*)+*p*_*IR*_(*t*) = 1.

Let *N*_*ab*_(*t*) denote the number of individuals observed to move from state *a* to state *b* between days *t* and *t*+1, and let *N*_*a*_(*t*) =∑_*b∈A*(*a*)_ *N*_*ab*_(*t*) be the number of individuals at risk in state *a* on day *t*. Under (1)–(2), conditional on *N*_*a*_(*t*), the admissible transition counts follow a restricted multinomial model,

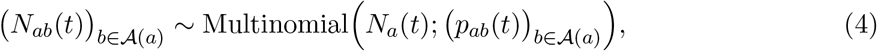

with likelihood contribution 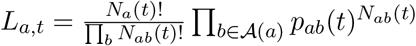.

#### Proposition 3.1

(Constrained MLE). *For N*_*a*_(*t*) *>* 0, *a ∈ {S, E, I}, and b ∈ A*(*a*), *the maximum likelihood estimator is*

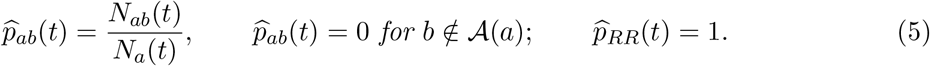

*Proof*. Maximizing *ℓ*_*a,t*_ = ∑_*b∈A*(*a*)_ *N*_*ab*_(*t*) log *p*_*ab*_(*t*) subject to ∑_*b∈A*(*a*)_ *p*_*ab*_(*t*) = 1 via a Lagrange multiplier *λ* gives the first order condition *N*_*ab*_(*t*)*/p*_*ab*_(*t*)+*λ* = 0, so *p*_*ab*_(*t*) = *−N*_*ab*_(*t*)*/λ*; summing over *b ∈ A*(*a*) and imposing the constraint yields *λ* = *−N*_*a*_(*t*), hence (5). Strict concavity of *ℓ*_*a,t*_ on the open simplex confirms this is the unique maximizer.

### 3.3 Reduction to a Binomial Model and Finite Sample Efficiency

Because |*A*(*a*)| = 2 for each non-absorbing compartment *a ∈ {S, E, I}*, the multinomial in (4) degenerates to a binomial model. Writing *b*^*\**^ (*S*) = *E, b*^*\**^ (*E*) = *I, b* (*I*) = *R* for the forward transition and 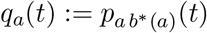, we have, conditional on *N*_*a*_(*t*),

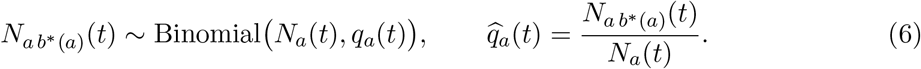

#### Proposition 3.2

(Efficiency). *The Fisher information for q*_*a*_(*t*) *is I*_*a*_(*t*) = *N*_*a*_(*t*)*/*[*q*_*a*_(*t*)(1 *− q*_*a*_(*t*))], *and* Var 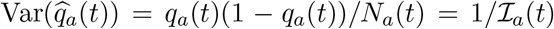 *exactly, for every finite N*_*a*_(*t*); *the constrained MLE is thus fully efficient, not merely asymptotically so*.

*Proof*. With 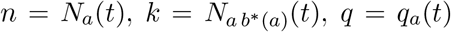, the log likelihood is *ℓ*(*q*) = *k* log *q* + (*n − k*) log(1 *− q*). The score *U* (*q*) = *k/q −* (*n − k*)*/*(1 *− q*) = (*k − nq*)*/*[*q*(1 *− q*)] vanishes at 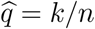. The negative expected curvature is *I*_*a*_(*t*) = *−*E[*∂*^2^*ℓ/∂q*^2^] = *n/q* + *n/*(1 *− q*) = *n/*[*q*(1 *− q*)]. Since *k* | *n ∼* Binomial(*n, q*), Var(*k* |*n*) = *nq*(1 *− q*) exactly, so Var 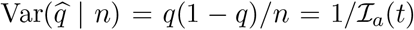, attaining the Cramér Rao bound with equality.

#### Proposition 3.3

(Unbiasedness and consistency). 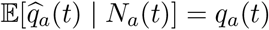, *and* 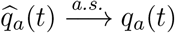*as N*_*a*_ *→ ∞* (*t*)*by the Strong Law of Large Numbers applied to the sum of i*.*i*.*d. Bernoulli*(*q*_*a*_(*t*)) *trials underlying* 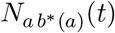.

### 3.4 Asymptotic Normality and Confidence Intervals

#### Theorem 3.4

(Asymptotic normality). 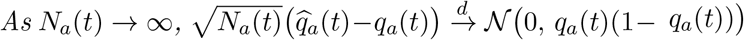

*Proof*. Conditional on *N*_*a*_(*t*) = *n*, the Markov property (1) implies 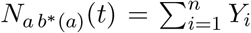 with 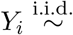 Bernoulli 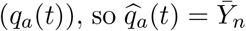. The Lindeberg Lévy central limit theorem gives 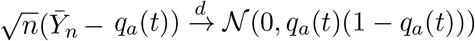 substituting *n* = *N*_*a*_(*t*) completes the proof.

A Wald interval follows immediately by Slutsky’s theorem, but performs poorly when *N*_*a*_(*t*) is small or *q*_*a*_(*t*) is close to a boundary precisely the regime relevant to sparsely reported district days. We therefore use the Wilson score interval (Wilson, 1927), obtained by inverting the score statistic 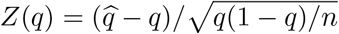 rather than the Wald statistic:

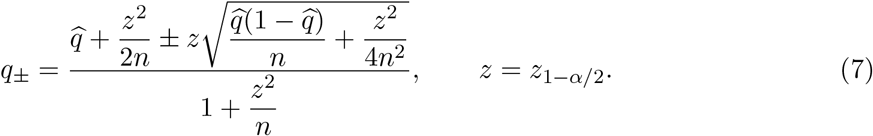

### 3.5 Temporal Smoothing and a Bandwidth Goodness of Fit Diagnostic

Because *N*_*a*_(*t*) is small on many district days, we stabilize 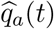 via a triangular kernel smoother of bandwidth *h*, analogous to classical kernel smoothing methods (Silverman, 1986) and to the non-parametric transition-matrix estimator of Aalen and Johansen (1978) for non-homogeneous Markov chains. With weights *w*_*h*_(*u*) = max(0, 1 *−* |*u*|*/h*),

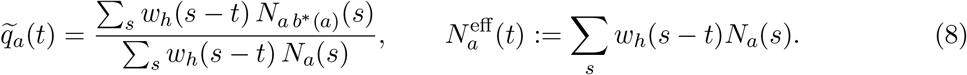

Under local stationarity of *q*_*a*_ (*·*), Var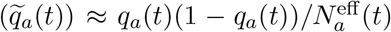, so that larger *h* trades reduced variance for potential bias. We select among candidate bandwidths using an aggregate Pearson goodness of fit statistic,

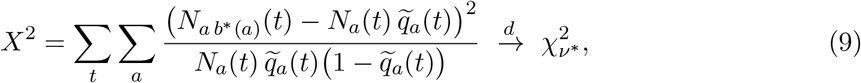

with *v*^*\**^ equal to the number of admissible binomial cells less the effective degrees of freedom consumed by the smoother; large *X*^2^ relative to 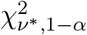 signals over-smoothing.

### 3.6 Likelihood Ratio Test for Time-Homogeneity

To formally test whether a given district and compartment genuinely require a time varying specification, we consider the nested hypotheses

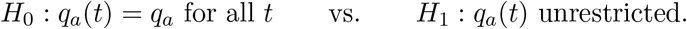

Under *H*_0_ the pooled MLE is 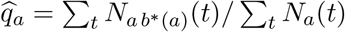, and the likelihood ratio statistic is

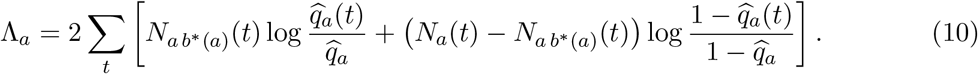

#### Proposition 3.5

*Under H*_0_, *as* 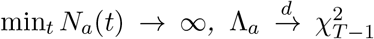, *by Wilks ‘ theorem (Wilks, 1938), since H*_0_ *is a one parameter nested restriction of the T -parameter model H*_1_.

We reject *H*_0_ concluding that time-inhomogeneity is statistically necessary at level *α* if 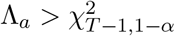

### 3.7 Hybrid Stochastic Machine Learning Extension

The purely non-parametric estimator 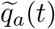 in (8) explains that *q*_*a*_(*t*) varies with time but not why. We therefore model the logit transformed transition probability as a function of the observable covariate vector

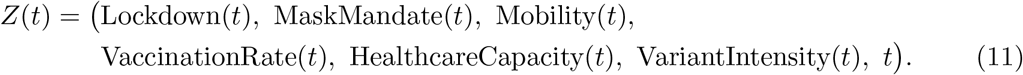

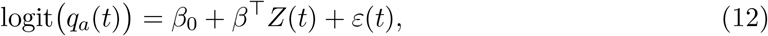

fitted as a weighted logistic generalized linear model with binomial denominators *N*_*a*_(*t*) the stochastic component). As a fully non-parametric alternative that does not impose the logit linear functional form, we fit a random forest (Breiman, 2001)

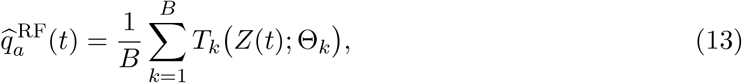

where *T*_*k*_(*·*; Θ_*k*_) is a regression tree grown on a bootstrap resample with independently randomized feature subsets Θ_*k*_, targeting the empirical proportion 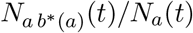 weighted by *N*_*a*_(*t*). Models (12) and (13) together with the homogeneous pooled baseline 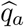 are compared out of sample using the weighted Brier score

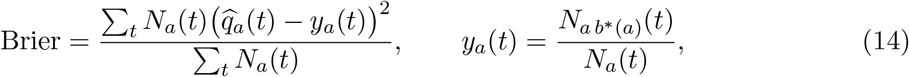

and the held out binomial log likelihood 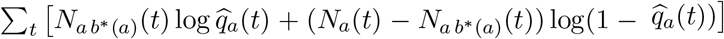.

## 4 Data Analysis and Reporting

### 4.1 Data

We use a compiled daily, district level COVID-19 surveillance panel for Sierra Leone covering 1 March 2020 to 31 December 2023 (1,401 days) across all 16 administrative districts (22,416 district day observations). For each district day the panel records laboratory confirmed cases, clinically suspected cases, deaths, recoveries, an estimate of currently active cases (cumulative confirmed minus cumulative recovered and deceased), an estimate of currently exposed contacts, hospitalizations, the vaccination rate, binary lockdown and mask-mandate indicators, a relative mobility index, a healthcare capacity index, and the dominant circulating SARS-CoV-2 variant with an associated transmissibility/severity intensity score.

### 4.2 SEIR Compartment Reconstruction

The raw surveillance panel reports daily flow and stock quantities rather than ready made transition counts, so we implement an explicit, documented reconstruction pipeline in R. For each district we treat the daily count of newly identified exposed contacts as the inflow *S → E*, the daily count of newly confirmed cases as the outflow *E → I*, and the sum of daily recoveries and deaths as the outflow *I → R*. The exposed compartment stock is reconstructed recursively,

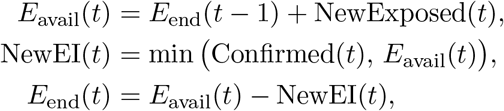

with *E*_end_(0) = 0, and *N*_*E*_(*t*) := *E*_avail_(*t*) is used as the at risk denominator for the *E*-row. The infectious compartment stock *N*_*I*_(*t*) is taken directly from the reported active case count, and the daily removal count is capped at *N*_*I*_(*t*) to respect the SEIR structural constraint. The removed compartment stock is obtained by simple accumulation, *R*(*t*) =_*s≤ t*_ New IR(*s*), which requires no further assumptions since it is a pure running total of observed daily recoveries and deaths. This bookkeeping construction inferring compartment occupancy from reported incidence, recovery, and mortality flows follows standard practice in surveillance-based compartmental reconstruction and mirrors the logic of the non-parametric transition matrix estimator of Aalen and Johansen (1978).

Because the panel does not report a susceptible population denominator, we cannot construct a bounded at risk count *N*_*S*_(*t*) for the district population and therefore cannot estimate *p*_*SE*_(*t*) as a constrained MLE in the same sense as *p*_*EI*_(*t*) and *p*_*IR*_(*t*). Rather than substitute an external, and for several of the 16 current districts internally inconsistent, census figure (the 2017 creation of Karene and Falaba districts from portions of Bombali, Port Loko, Kambia, and Koinadugu makes population reapportionment to current district boundaries non-trivial and, in the sources we consulted, inconsistent), we instead report the *S → E* boundary as an observed daily *incidence flow* the number of newly identified exposed contacts per day, NewExposed(*t*) smoothed with the same triangular kernel as (8). This flow is descriptively informative about the timing and intensity of new transmission even though, unlike *p*_*EI*_(*t*) and *p*_*IR*_(*t*), it is not a probability bounded in [0, 1] and does not carry a Wilson confidence interval or a likelihood-ratio test of homogeneity under the framework of Section 3. We restrict formal statistical estimation, testing, and the hybrid model of Section 3.6 to the empirically well defined *E* row and *I* row sub-chain, and return to this scope limitation in Section 6.

### 4.3 Estimation and Testing Pipeline (implemented in R)

All computation was carried out in R (with `dplyr, ggplot2, and randomForest`). For each district and each of the two estimable rows we compute: (i) the raw daily MLE 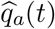 from (5); (ii) the triangular kernel smoothed estimator 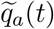 from (8) with bandwidth *h* = 14 days, together with 95% Wilson score intervals (7); (iii) the likelihood ratio statistic Λ_*a*_ from (10) testing *H*_0_: time-homogeneity against *H*_1_: unrestricted time variation; and (iv) the aggregate goodness of fit statistic (9) evaluated at four candidate bandwidths (*h* ∈ {7, 14, 28, 60}days) to assess whether *h* = 14 over or under smooths relative to the raw counts.

For the hybrid stochastic machine learning comparison (Section 3.6), we restrict to district days with *N*_*E*_(*t*) *≥* 5 (to ensure a minimally informative binomial trial size), split the resulting panel temporally into a training set (all observations before 1 January 2023) and a held out test set (all of 2023), and fit: (a) the time-homogeneous pooled baseline *q*_*E*_; (b) a weighted logistic GLM (12) with covariates Lockdown, Mask Mandate, Mobility Level, Vaccination Rate, Healthcare Capacity, Variant Intensity, and a linear time index; and (c) a random forest (13) with 400 trees and mtry = 3 on the same covariates. All three models are evaluated out of sample on the 2023 test set using the weighted Brier score (14) and the held out binomial log likelihood.

Finally, to present the complete SEIR picture rather than the *E* and *I* rows in isolation, we additionally compute, per district: the smoothed *S → E* incidence flow NewExposed(*t*); the exposed compartment stock *E*_end_(*t*); the infectious compartment stock *N*_*I*_(*t*); and the cumulative removed compartment stock *R*(*t*). These four series are visualized together (Section 5.5) to give a full compartment level view of the epidemic alongside the two rigorously estimated transition probabilities. The complete R pipeline comprises four scripts: `01 data processing.R` (compartment reconstruction), `02 modeling.R` (kernel smoothing, Wilson intervals, the likelihood ratio test, the goodness of fit diagnostic, and the hybrid GLM/random forest comparison), `03 figures.R` (Figures 1–7), and `04 full SEIR figures.R` (Figures 8–12); full source code is reproduced in Appendix A.

**Figure 1:**
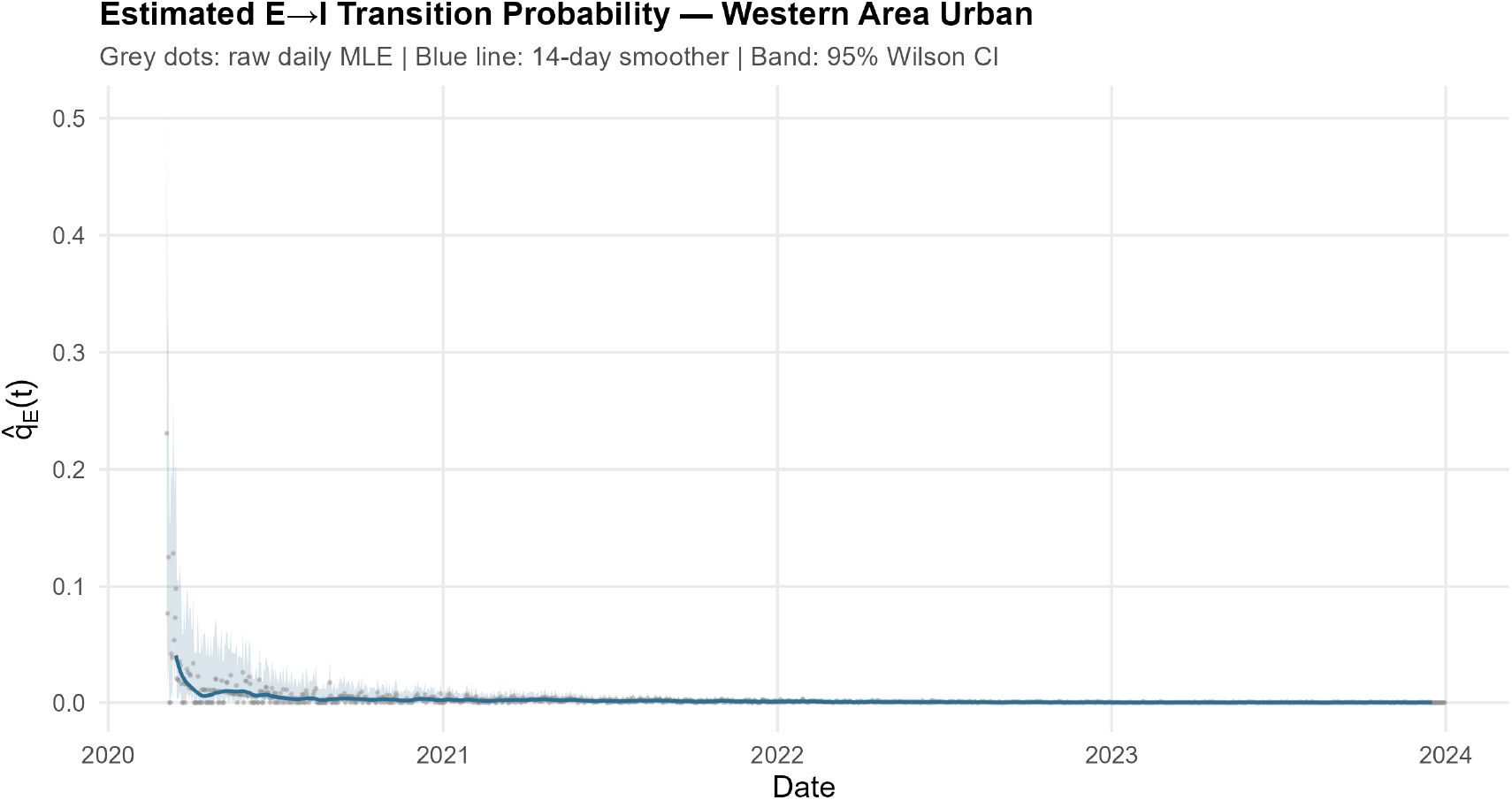
Estimated exposed to infectious transition probability 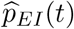 for Western Area Urban. Grey points: raw daily constrained MLE (Eq. 5); blue line: 14-day triangular kernel smoothed estimate (Eq. 8); shaded band: 95% Wilson score interval (Eq. 7).

**Figure 2:**
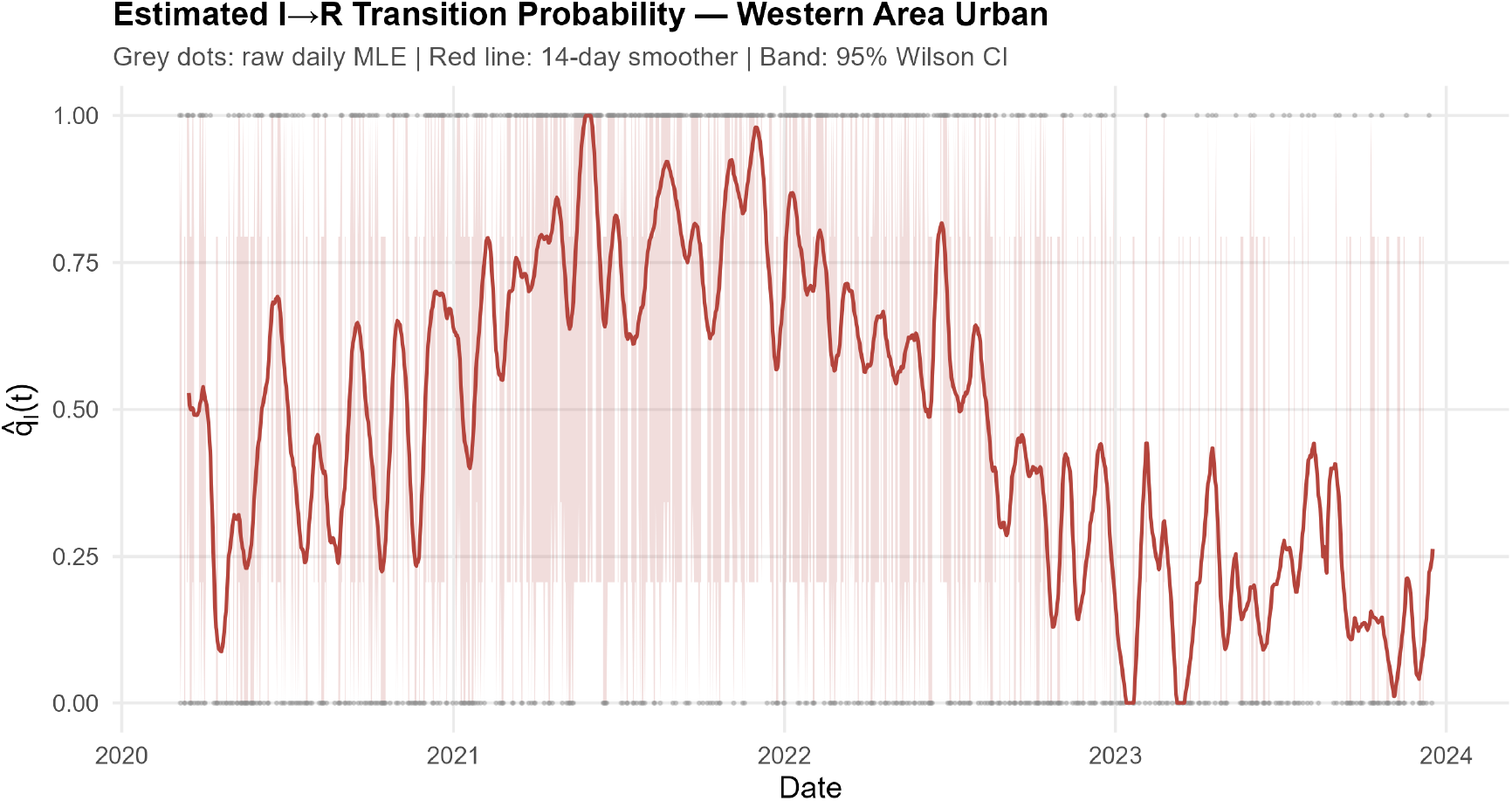
Estimated infectious to removed transition probability *p*_*IR*_(*t*) for Western Area Urban, with the same conventions as Figure 1.

**Figure 3:**
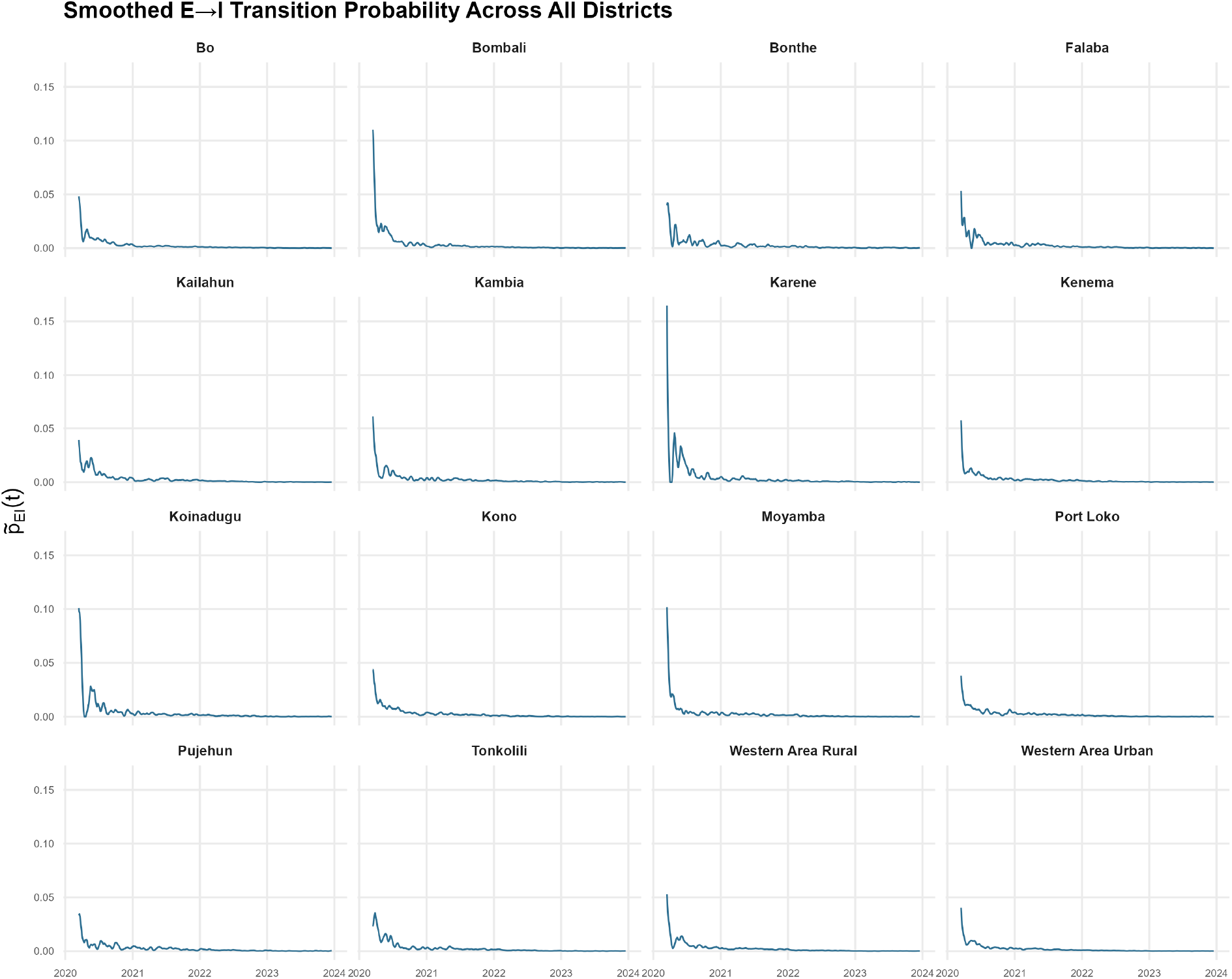
Smoothed exposed to infectious transition probability 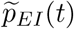 across all 16 districts, 2020–2023.

**Figure 4:**
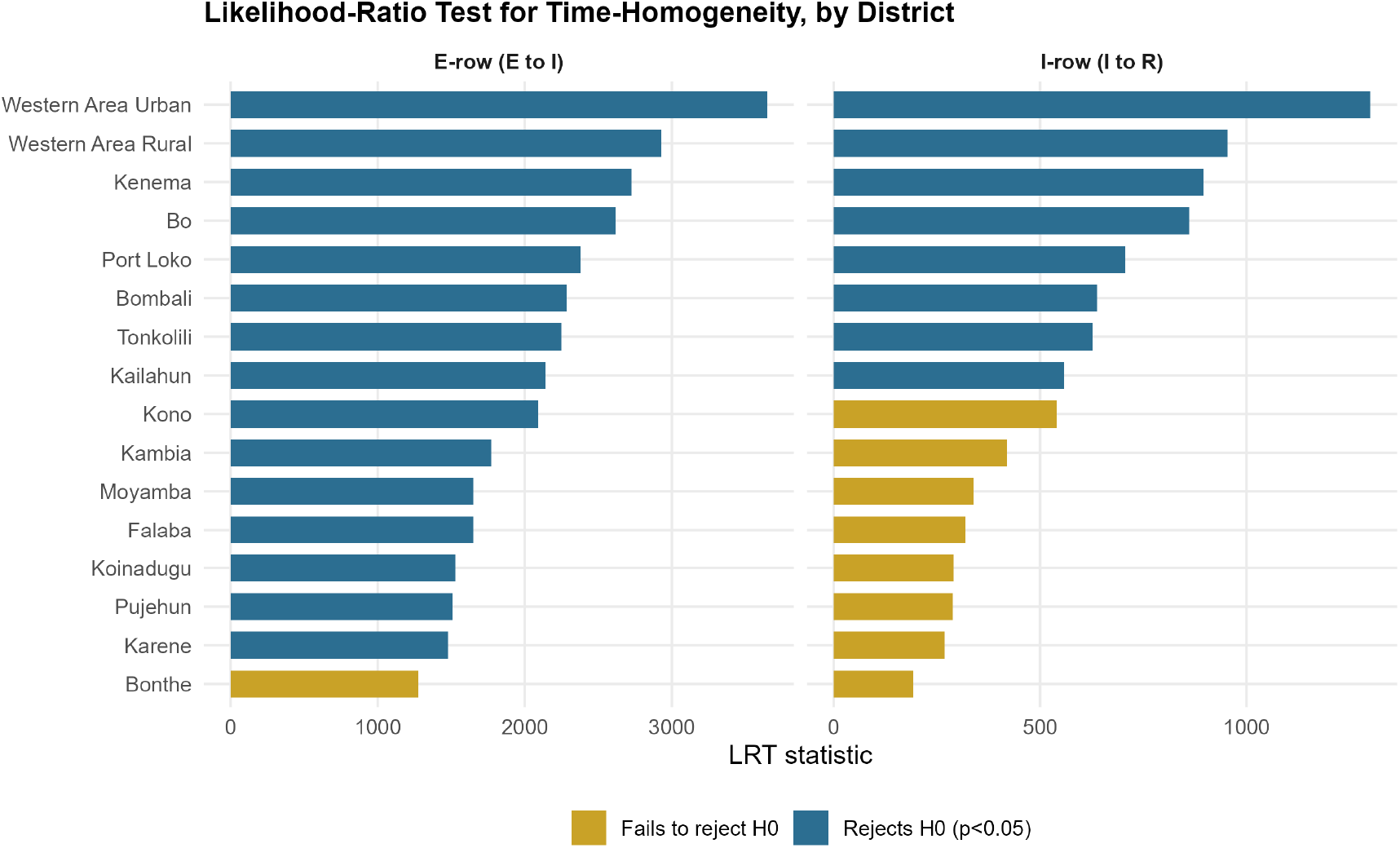
Likelihood ratio statistic Λ_*a*_ testing *H*_0_ : *q*_*a*_(*t*) = *q*_*a*_ against *H*_1_: unrestricted *q*_*a*_(*t*), by district and row. Blue bars: districts rejecting *H*_0_ at *α* = 0.05; gold bars: districts failing to reject *H*_0_.

**Figure 5:**
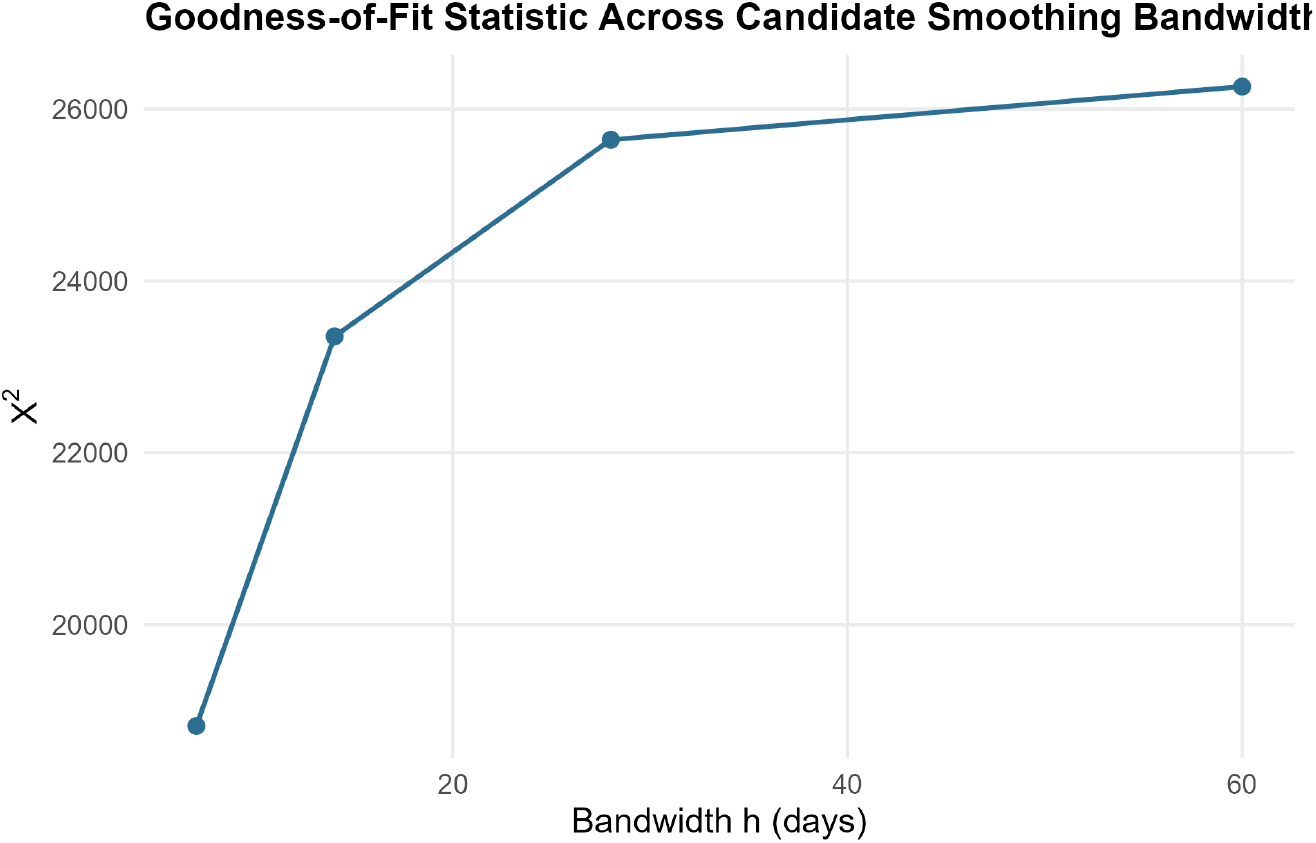
Goodness of fit statistic *X*^2^ (Eq. 9) as a function of candidate bandwidth *h*.

**Figure 6:**
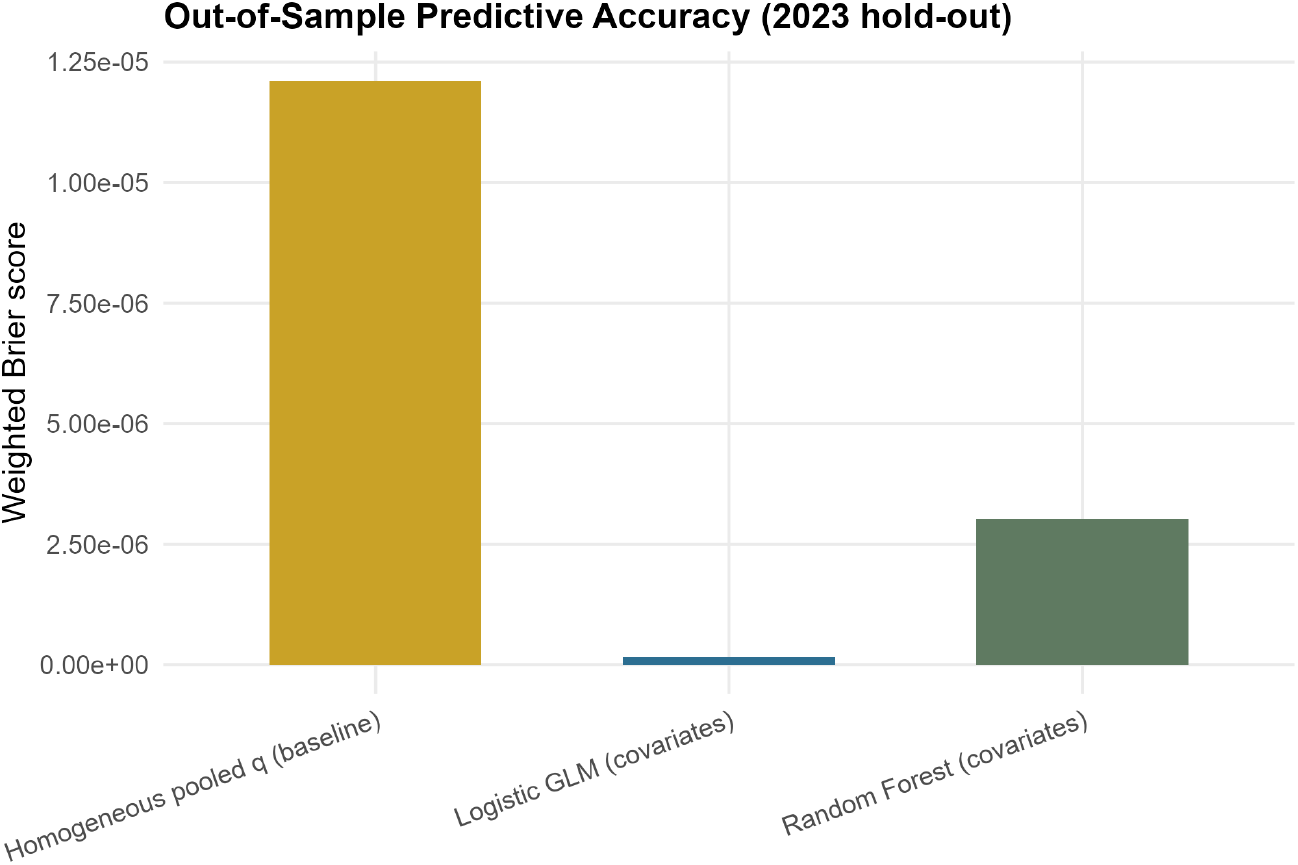
Out-of-sample weighted Brier score (Eq. 14) for the three competing models of *p*_*EI*_(*t*) (lower is better).

**Figure 7:**
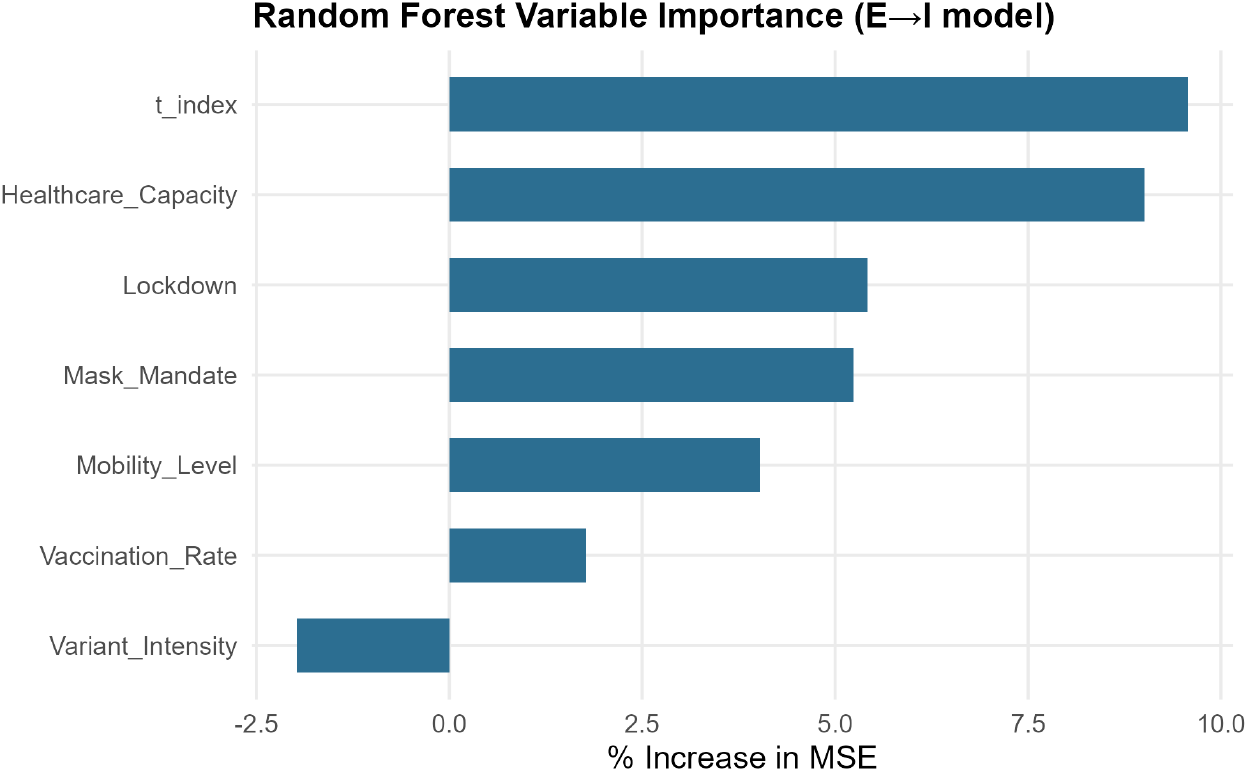
Random forest permutation variable importance for the *E → I* transition probability model.

**Figure 8:**
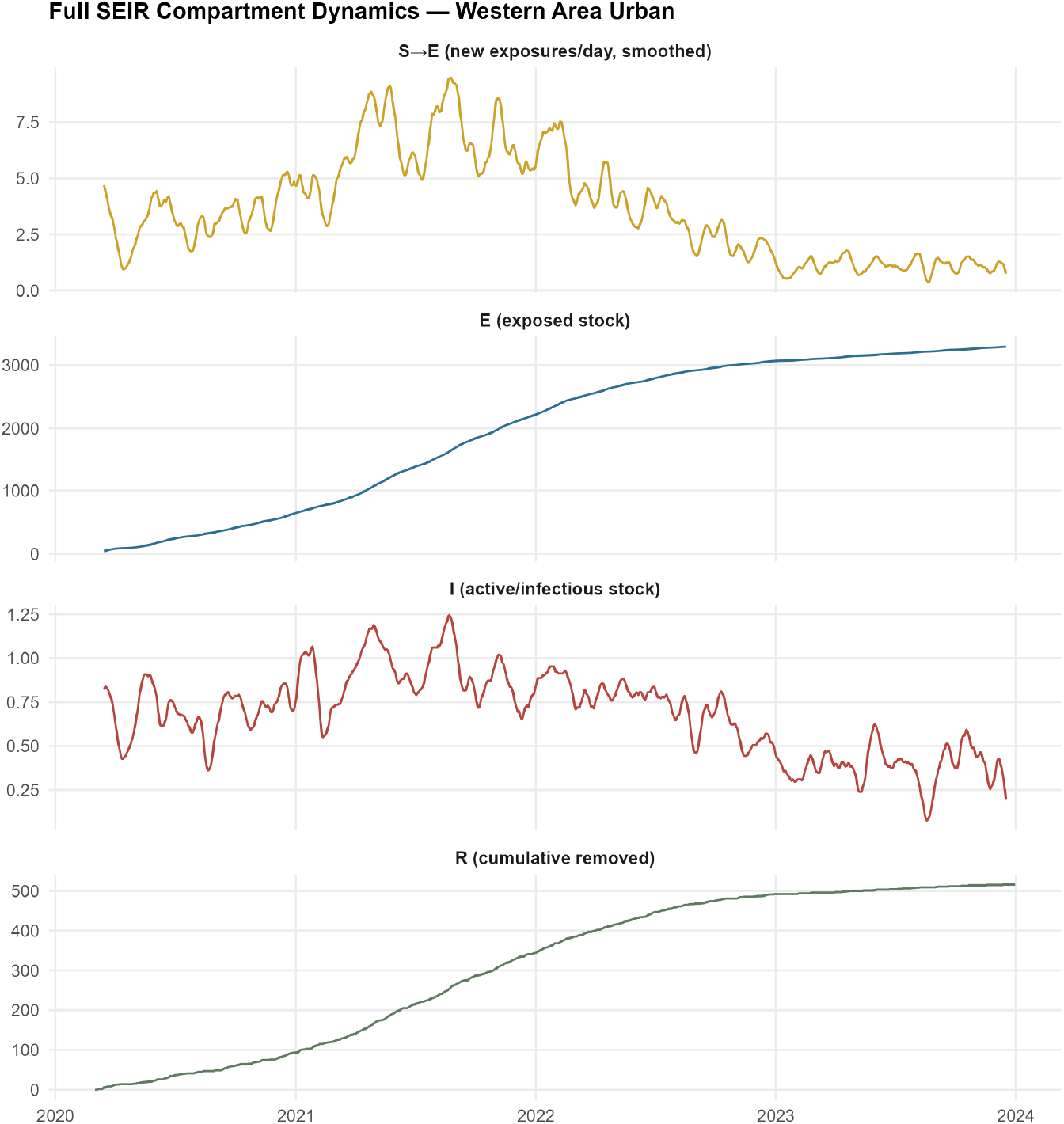
Full SEIR compartment dynamics for Western Area Urban: smoothed *S → E* incidence flow, exposed compartment stock *E* (*t*), infectious compartment stock *I*(*t*), and cumulative removed compartment stock *R*(*t*), 2020–2023.

**Figure 9:**
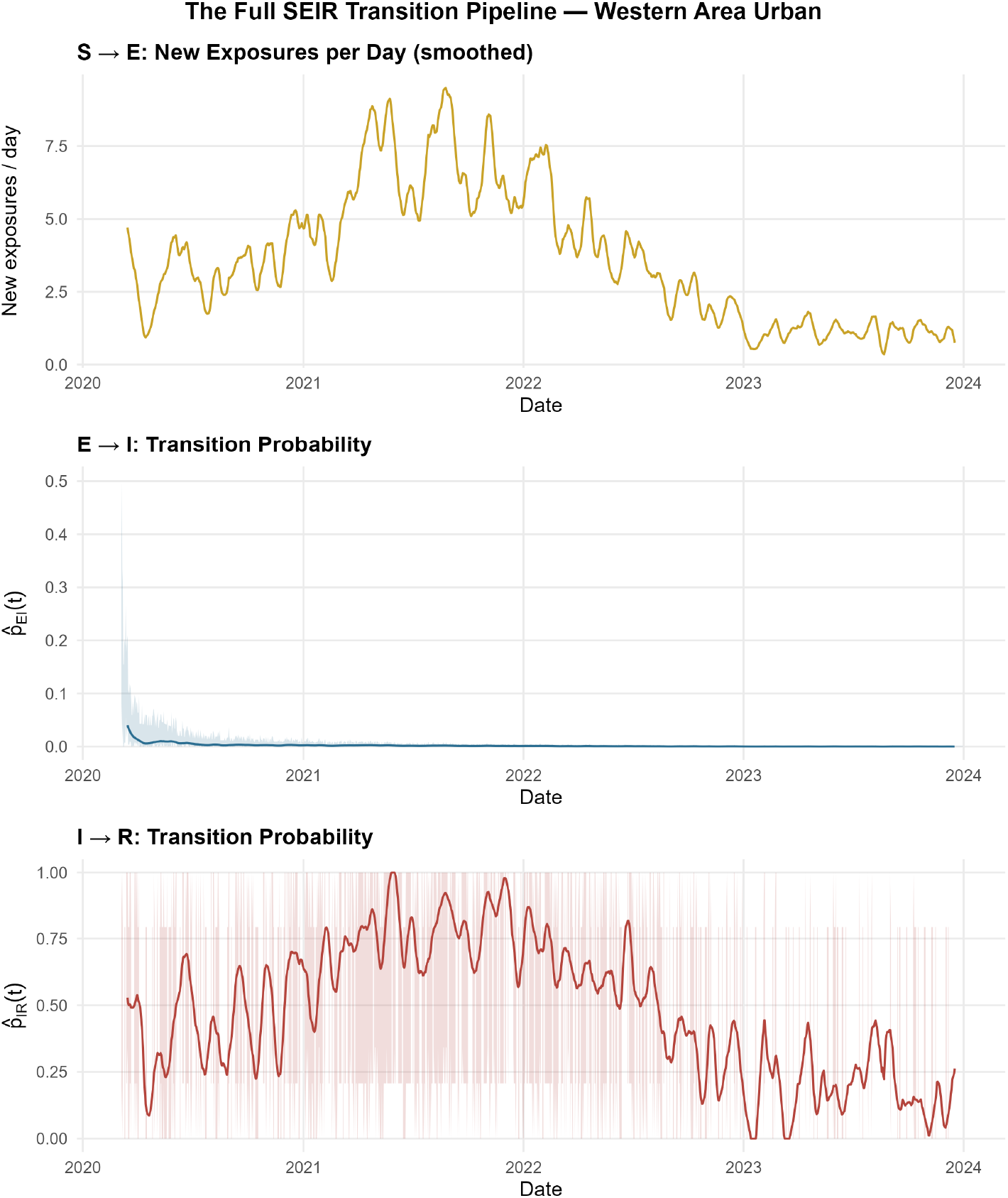
The full SEIR transition pipeline for Western Area Urban: smoothed *S → E* incidence (top, count scale), smoothed *E → I* transition probability with 95% Wilson band (middle), and smoothed *I → R* transition probability with 95% Wilson band (bottom).

**Figure 10:**
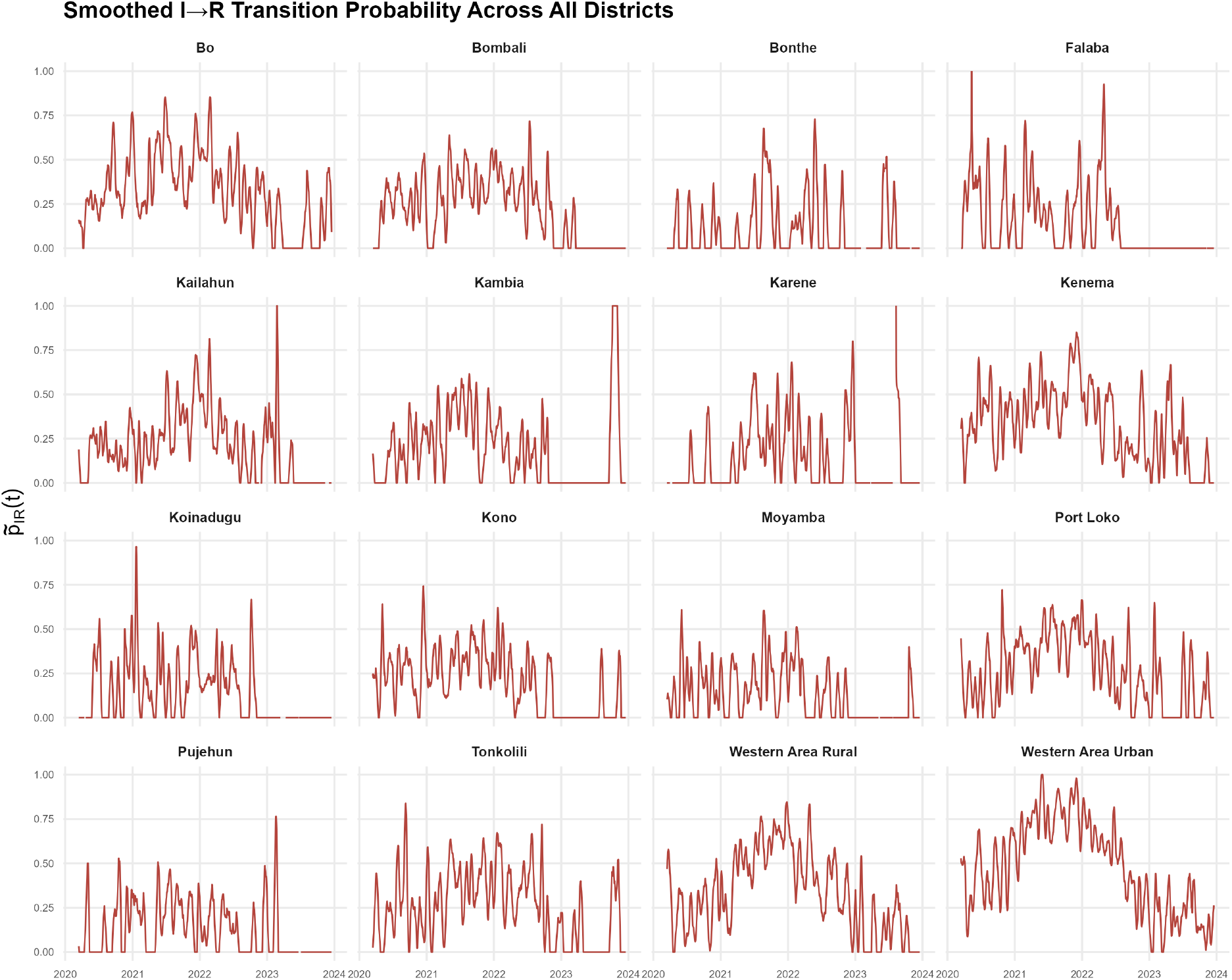
Smoothed *I → R* transition probability across all 16 districts, 2020–2023.

**Figure 11:**
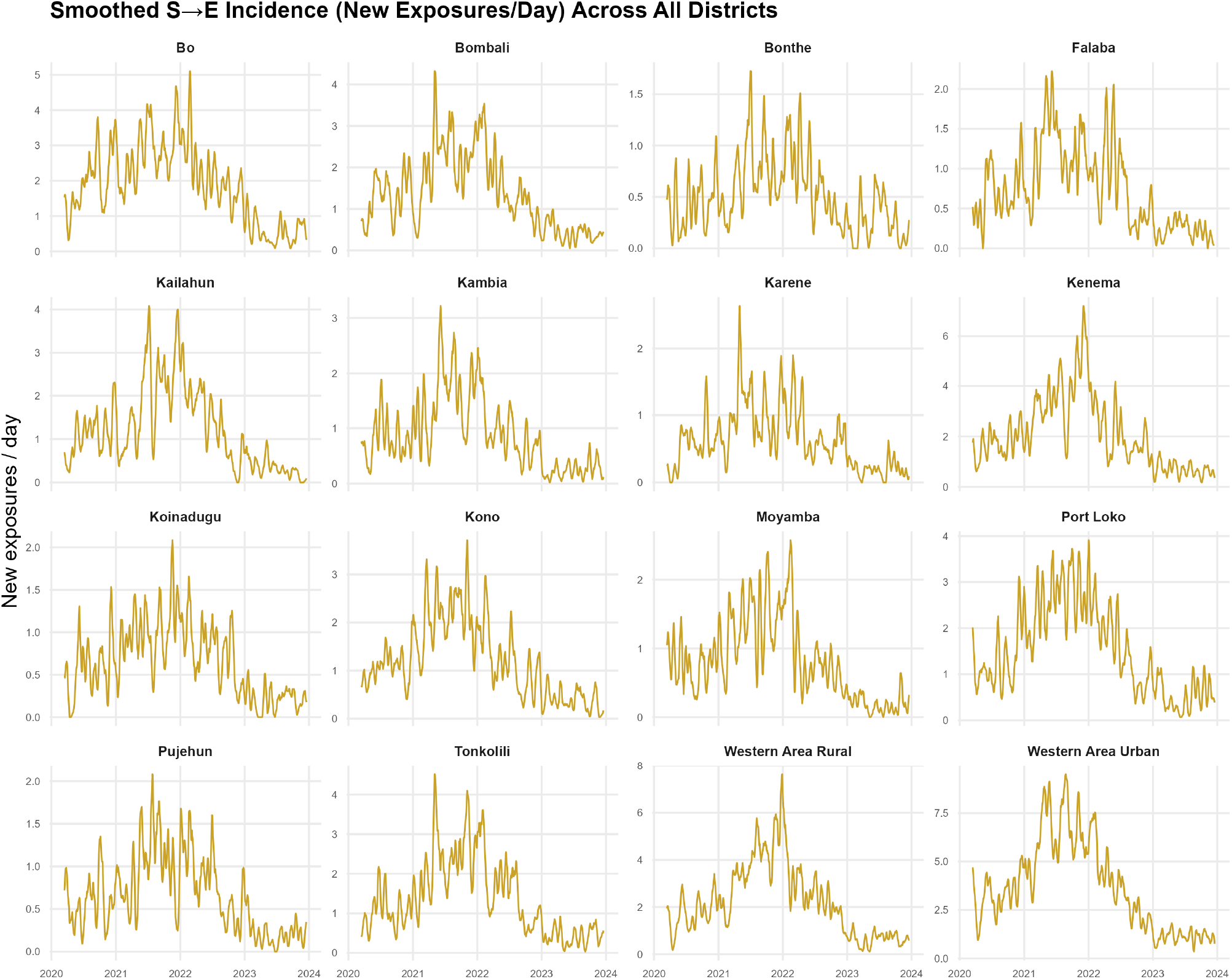
Smoothed *S → E* incidence flow (new exposures per day) across all 16 districts, 2020–2023. Note the free *y*-axis scale per panel, reflecting the wide range of district caseloads.

**Figure 12:**
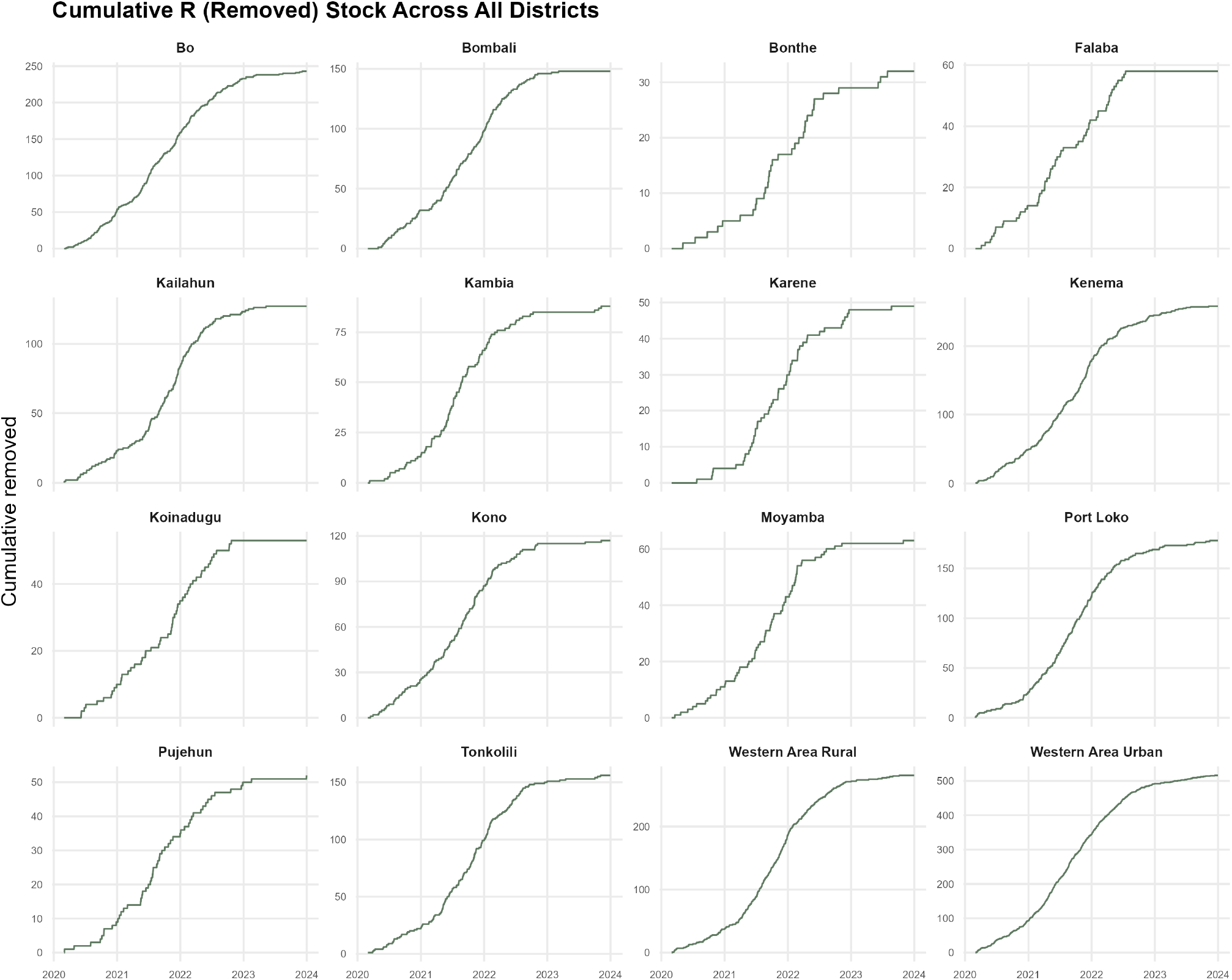
Cumulative removed compartment stock *R*(*t*) (recovered plus deceased) across all 16 districts, 2020–2023.

## 5 Results

### 5.1 Time Varying Transition Probabilities

Figure 1 shows the raw and 14 day kernel smoothed estimate of the exposed to infectious transition probability 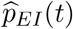 for Western Area Urban (the capital district and largest contributor to the national caseload), together with its 95% Wilson confidence band; Figure 2 shows the corresponding infectious to removed probability 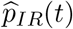. Both series exhibit pronounced, sustained departures from any constant level, with 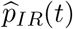 in particular showing multi month regimes of markedly different removal (recovery/death) probability consistent with successive epidemic waves. Figure 3 presents the smoothed *p*_*EI*_(*t*) curves for all 16 districts simultaneously, illustrating that the timing and magnitude of these regime shifts differ materially across districts.

### 5.2 Likelihood Ratio Test for Time-Homogeneity

Table 1 and Figure 4 report the likelihood ratio statistic Λ_*a*_ and associated *p* value for each district and each of the two estimable rows. At the *α* = 0.05 level, *H*_0_ (time-homogeneity) is rejected for the *E* row in 15 of 16 districts (all except Bonthe, *p* = 0.992) and for the *I* row in 8 of 16 districts. The rejections are overwhelming in magnitude for the most populous districts: Λ_*E*_ = 3647.5 for Western Area Urban and Λ_*E*_ = 2930.6 for Western Area Rural, both with *p <* 10^*−*100^ on *T −* 1 *≈* 1396–1399 degrees of freedom. The *I* row rejections are concentrated in districts with the largest cumulative caseloads (Western Area Urban, Western Area Rural, Bo, Kenema, Port Loko, Tonkolili), while several predominantly rural districts with smaller and sparser active case counts (Bonthe, Falaba, Karene, Koinadugu, Moyamba, Pujehun) do not reject time-homogeneity for the removal probability, plausibly reflecting lower statistical power in districts with fewer active case days rather than genuinely constant removal dynamics.

**Table 1:**
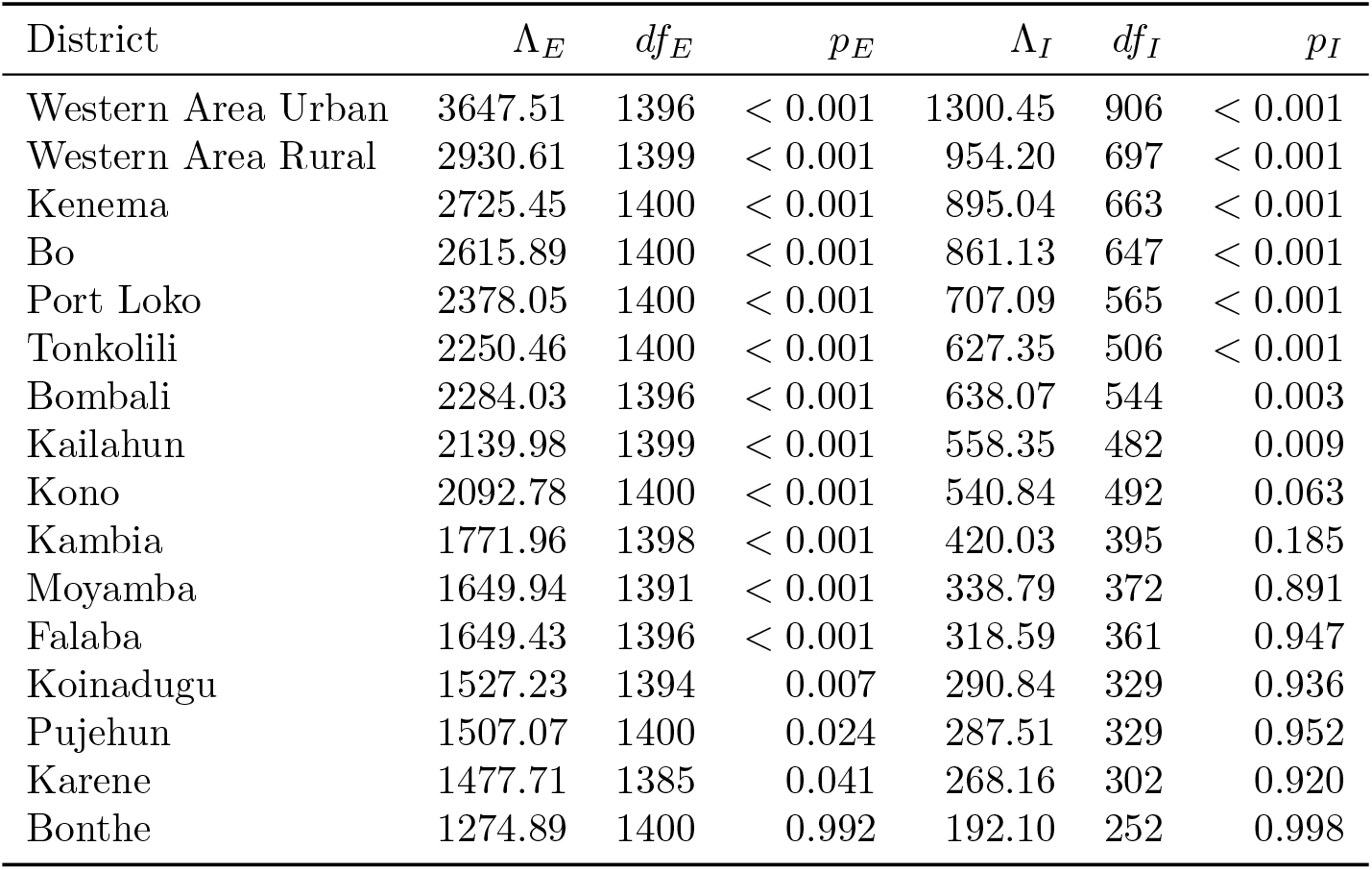
Likelihood ratio test of time-homogeneity, by district. Λ_*E*_, Λ_*I*_: LRT statistics for the*E → I* and *I → R* rows; *p*_*E*_, *p*_*I*_: corresponding *p* values against 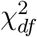.

| District | $\Lambda_E$ | $df_E$ | $p_E$ | $\Lambda_I$ | $df_I$ | $p_I$ |
| --- | --- | --- | --- | --- | --- | --- |
| Western Area Urban | 3647.51 | 1396 | < 0.001 | 1300.45 | 906 | < 0.001 |
| Western Area Rural | 2930.61 | 1399 | < 0.001 | 954.20 | 697 | < 0.001 |
| Kenema | 2725.45 | 1400 | < 0.001 | 895.04 | 663 | < 0.001 |
| Bo | 2615.89 | 1400 | < 0.001 | 861.13 | 647 | < 0.001 |
| Port Loko | 2378.05 | 1400 | < 0.001 | 707.09 | 565 | < 0.001 |
| Tonkolili | 2250.46 | 1400 | < 0.001 | 627.35 | 506 | < 0.001 |
| Bombali | 2284.03 | 1396 | < 0.001 | 638.07 | 544 | 0.003 |
| Kailahun | 2139.98 | 1399 | < 0.001 | 558.35 | 482 | 0.009 |
| Kono | 2092.78 | 1400 | < 0.001 | 540.84 | 492 | 0.063 |
| Kambia | 1771.96 | 1398 | < 0.001 | 420.03 | 395 | 0.185 |
| Moyamba | 1649.94 | 1391 | < 0.001 | 338.79 | 372 | 0.891 |
| Falaba | 1649.43 | 1396 | < 0.001 | 318.59 | 361 | 0.947 |
| Koinadugu | 1527.23 | 1394 | 0.007 | 290.84 | 329 | 0.936 |
| Pujehun | 1507.07 | 1400 | 0.024 | 287.51 | 329 | 0.952 |
| Karene | 1477.71 | 1385 | 0.041 | 268.16 | 302 | 0.920 |
| Bonthe | 1274.89 | 1400 | 0.992 | 192.10 | 252 | 0.998 |

### 5.3 Bandwidth Goodness of Fit

Table 2 reports the aggregate goodness of fit statistic (9) evaluated at four candidate bandwidths. All four bandwidths yield *p* ≈ 1, indicating no detectable lack of fit even at the widest window tested (*h* = 60 days); Figure 5 shows that *X*^2^ increases only gradually with *h*. We adopt *h* = 14 days for the substantive results above as a bandwidth that materially reduces the variance of the raw daily estimator (Figures 1–2) while remaining well within the region of adequate fit.

**Table 2:**
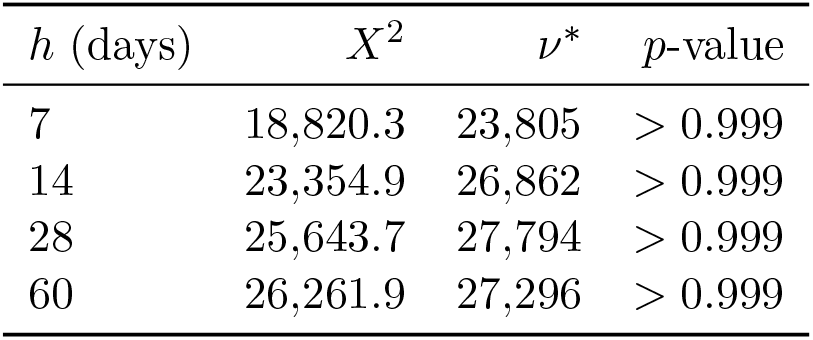
Aggregate goodness of fit statistic across candidate smoothing bandwidths (pooled across all 16 districts).

| $h$ (days) | $X^2$ | $\nu^*$ | $p$-value |
| --- | --- | --- | --- |
| 7 | 18,820.3 | 23,805 | $> 0.999$ |
| 14 | 23,354.9 | 26,862 | $> 0.999$ |
| 28 | 25,643.7 | 27,794 | $> 0.999$ |
| 60 | 26,261.9 | 27,296 | $> 0.999$ |

### 5.4 Hybrid Stochastic Machine Learning Comparison

Table 3 reports the out of sample (2023 hold out) predictive performance of the time-homogeneous pooled baseline, the covariate augmented logistic GLM, and the random forest, for the *E → I* transition probability. The logistic GLM achieves the lowest weighted Brier score (1.60 *×* 10^*−*7^, versus 1.21 *×* 10^*−*5^ for the homogeneous baseline a reduction by a factor of approximately 76 and 3.97 *×* 10^*−*6^ for the random forest) and the highest held out log likelihood, indicating that a parsimonious, logit linear combination of policy and mobility covariates captures the dominant time-inhomogeneity in *p*_*EI*_(*t*) more effectively than either assuming homogeneity or fitting a fully non-parametric ensemble on the same covariate set. Figure 6 displays the three Brier scores; Figure 7 shows the random forest permutation variable importance ranking, in which healthcare capacity and the linear time index are the two most influential predictors, followed by mobility level, lockdown status, and mask mandate status, while the vaccination rate and variant intensity score contribute comparatively little marginal predictive value in the *E → I* model.

**Table 3:** Out of sample (2023 hold out) predictive performance for the *E → I* transition probability.

| Model | Weighted Brier score | Held out log likelihood |
| --- | --- | --- |
| Homogeneous pooled $\hat{q}_E$ (baseline) | $1.21 \times 10^{-5}$ | -29,048.7 |
| Logistic GLM (covariates) | $1.60 \times 10^{-7}$ | -10,250.8 |
| Random forest (covariates) | $3.97 \times 10^{-6}$ | -19,723.4 |

### 5.5 Full SEIR Compartment Dynamics

The results above (Sections 5.1–5.4) concern the *E* row and *I* row transition probabilities, which are the only rows estimable as bounded probabilities without a district population denominator (Section 4.2). For completeness, Figure 8 presents the full compartment picture for Western Area Urban: the smoothed *S → E* incidence flow (new exposures per day), the exposed compartment stock *E*(*t*), the infectious compartment stock *I*(*t*), and the cumulative removed compartment stock *R*(*t*). The *S → E* incidence and *I*(*t*) stock both show clear multi-wave structure, with incidence peaking in mid to late 2021, while *E*(*t*) and *R*(*t*) being cumulative or near cumulative quantities rise monotonically and level off as the district’s compiled epidemic activity subsides after 2022. Figure 9 places the *S → E* incidence flow directly above the *E → I* and *I → R* smoothed transition probabilities for the same district, visually connecting the full compartmental chain: note that the *S → E* panel is plotted on an incidence count scale rather than a probability scale, since (as discussed in Section 4.2) no population based at risk denominator is available to bound it in [0, 1].

Figures 10–12 extend the all district small multiples view of Figure 3 to the remaining compartments: Figure 10 shows the smoothed *I → R* probability for all 16 districts, Figure 11 shows the smoothed *S → E* incidence flow for all 16 districts, and Figure 12 shows the cumulative *R* stock for all 16 districts. Read together with Figure 3, these four panels give a complete, district resolved picture of the entire SEIR chain underlying the transition probability estimates in Sections 5.1–5.4.

## 6 Discussion

The empirical results provide clear statistical evidence against the time-homogeneity assumption implicit in classical compartmental models of COVID-19 transmission in Sierra Leone. The near universal rejection of homogeneity for the exposed to infectious transition (15 of 16 districts) is consistent with the succession of variants of concern and shifting non-pharmaceutical interventions documented for Sierra Leone over the study period (Liu et al., 2022; Lin et al., 2022), and with the general finding that mobility sensitive Markov models outperform time-homogeneous alternatives elsewhere (Oka et al., 2020). The more mixed picture for the infectious to removed transition rejected in the eight districts with the largest cumulative caseloads but not in eight smaller, more rural districts is at least partly a power phenomenon: the likelihood ratio test in Proposition 3.7 requires a reasonably large number of active case days to distinguish genuine time variation from sampling noise, and the districts that fail to reject *H*_0_ are precisely those with the fewest active case days (*n*_*I*_ ranging from 252 to 373, versus up to 907 in Western Area Urban). This pattern illustrates a broader methodological point for sub-national surveillance in resource constrained settings: statistical power to detect time-inhomogeneity is itself heterogeneous across administrative units, and null results for smaller districts should not be over interpreted as evidence of genuinely constant transmission dynamics.

The hybrid stochastic machine learning comparison offers a complementary, and in one respect surprising, conclusion: the interpretable, logit-linear generalized linear model outperforms the random forest out-of-sample, despite the latter’s greater flexibility. We interpret this as evidence that the dominant source of time-inhomogeneity in *p*_*EI*_(*t*) over the study period is well captured by a smooth, monotonic combination of the available policy and mobility covariates together with a slow secular trend, so that the additional flexibility of a tree ensemble mainly fits noise in the 2020–2022 training period rather than generalizable signal, a pattern also noted in other COVID-19 forecasting comparisons between parametric and ensemble methods (Zhang et al., 2023). The random forest variable importance ranking is nonetheless informative in its own right: healthcare capacity and the underlying time trend dominate, ahead of mobility and policy indicators, suggesting that health system strain itself correlated with, but not identical to, the policy variables may be a more proximate driver of the estimated exposed to infectious probability than the policy indicators considered in isolation.

The full compartment view in Section 5.5 makes the scope of the estimable sub-chain concrete: the *S → E* boundary (Figures 8, 9, and 11) tracks new exposure incidence closely and shares the same multi-wave timing as the *I* (*t*) stock, which is consistent with the *E*-row and *I* row LRT rejections in Table 1 the incidence process driving new exposures is visibly non-stationary, so it would be surprising if the downstream *E → I* and *I → R* probabilities were constant. We nonetheless stop short of converting the *S → E* incidence flow into a bounded probability, since doing so would require dividing by a susceptible population count that is not available at the current (post-2017, 16-district) administrative resolution without reconciling inconsistent population reapportionments for the two districts (Karene and Falaba) created from portions of three other districts in 2017; we regard an approximate or interpolated population denominator as more likely to mislead than to inform, and prefer to report the incidence flow honestly as a count rather than dress it as a probability it is not.

Several limitations should be borne in mind. First, the compartment occupancy reconstruction in Section 4.2 necessarily makes assumptions about how routinely reported incidence, recovery, and mortality counts map onto SEIR compartment stocks and flows; while we believe the resulting pipeline is a reasonable and transparent approximation, direct contact tracing or line list data would permit more precise reconstruction of the exposed compartment stock in particular. Second, in the absence of a susceptible-population denominator in the surveillance panel, we could not estimate the *S → E* row within the same constrained likelihood framework, and future work should incorporate district-level population figures ideally sourced directly from Statistics Sierra Leone at the current 16-district resolution to complete the four state chain and extend the likelihood ratio and confidence interval machinery of Section 3 to *p*_*SE*_(*t*) as well. Third, the likelihood ratio test in Section 3.7 assumes independence of daily transitions conditional on the state occupied, which is a standard simplifying assumption in this literature but may understate serial correlation in reporting; a block bootstrap or sandwich corrected variance would be a natural robustness check. Finally, while the goodness of fit diagnostic in Section 3.5 did not detect lack of fit at any of the four bandwidths considered, this diagnostic evaluates fit to the (already smoothed) triangular kernel and cannot rule out other forms of misspecification, such as an incorrectly specified SEIR structural zero pattern in settings with substantial re-infection.

## 7 Conclusion

This paper developed a complete constrained likelihood and asymptotic inference framework for time-inhomogeneous SEIR Markov chains, encompassing the constrained maximum likelihood estimator, its finite sample efficiency and asymptotic normality, Wilson score confidence intervals, a likelihood ratio test of time-homogeneity, and a bandwidth selection goodness of fit diagnostic for the associated kernel smoothed estimator, and extended the framework into a stochastic machine learning hybrid by regressing the estimated transition probabilities on policy and mobility covariates via both a logistic generalized linear model and a random forest. Applied to a compiled daily, district level COVID-19 surveillance panel for Sierra Leone (16 districts, March 2020–December 2023), the likelihood ratio test rejected time-homogeneity of the exposed to infectious transition in 15 of 16 districts and of the infectious to removed transition in 8 of 16 districts, and the covariate augmented logistic model substantially outperformed both a time-homogeneous baseline and a random forest out of sample. These findings support the use of time-inhomogeneous, covariate informed Markov chain models over classical time-homogeneous compartmental models for sub-national COVID-19 surveillance in resource constrained settings, and they suggest that healthcare capacity strain and slow secular trends, more than the individual policy indicators considered in isolation, are the dominant correlates of estimated time variation in the exposed to infectious transition probability. Future work should extend the estimable state space to include the susceptible compartment using district level population denominators, incorporate serial correlation robust variance estimation for the likelihood ratio test, and evaluate the hybrid framework’s predictive performance prospectively on subsequent epidemic waves or other infectious diseases with a comparable SEIR structure.

## Data Availability

No primary data was generated by this study. The minimal surveillance dataset used in this study was derived from the Sierra Leone Ministry of Health and Sanitation / National Public Health Agency (NPHA) district-level COVID-19 surveillance reports. All processed data files and analytical code used to generate the results are included within the manuscript.

## A Reproducible R Implementation

This appendix reproduces, in full, the four R scripts used to process the data, fit and test the models of Section 3, generate Figures 1–12, and produce Tables 1–3. All computation was performed in R with the `dplyr, ggplot2, tidyr, randomForest`, and `patchwork` packages.

### A.1 01_data_processing.R SEIR Compartment Reconstruction

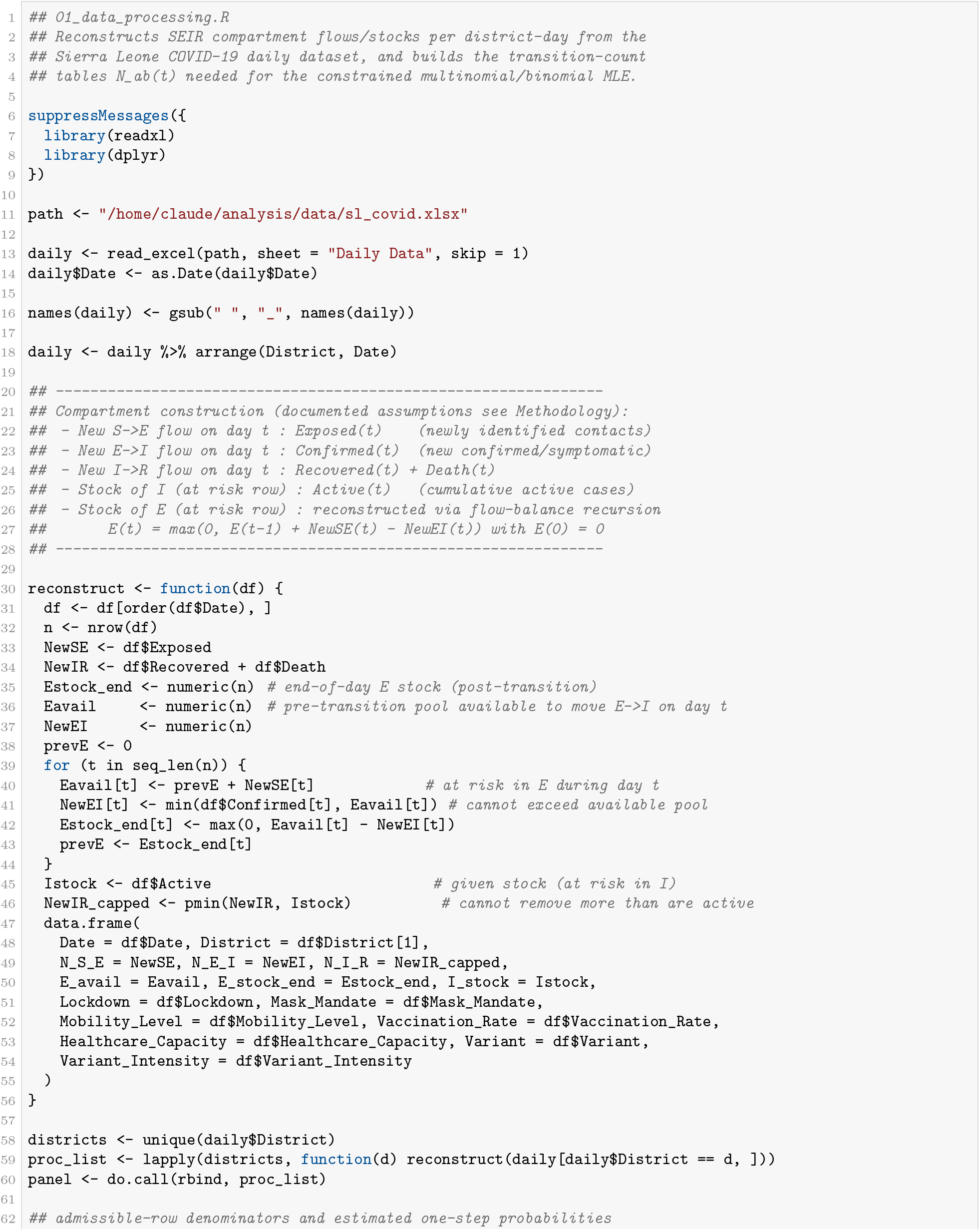
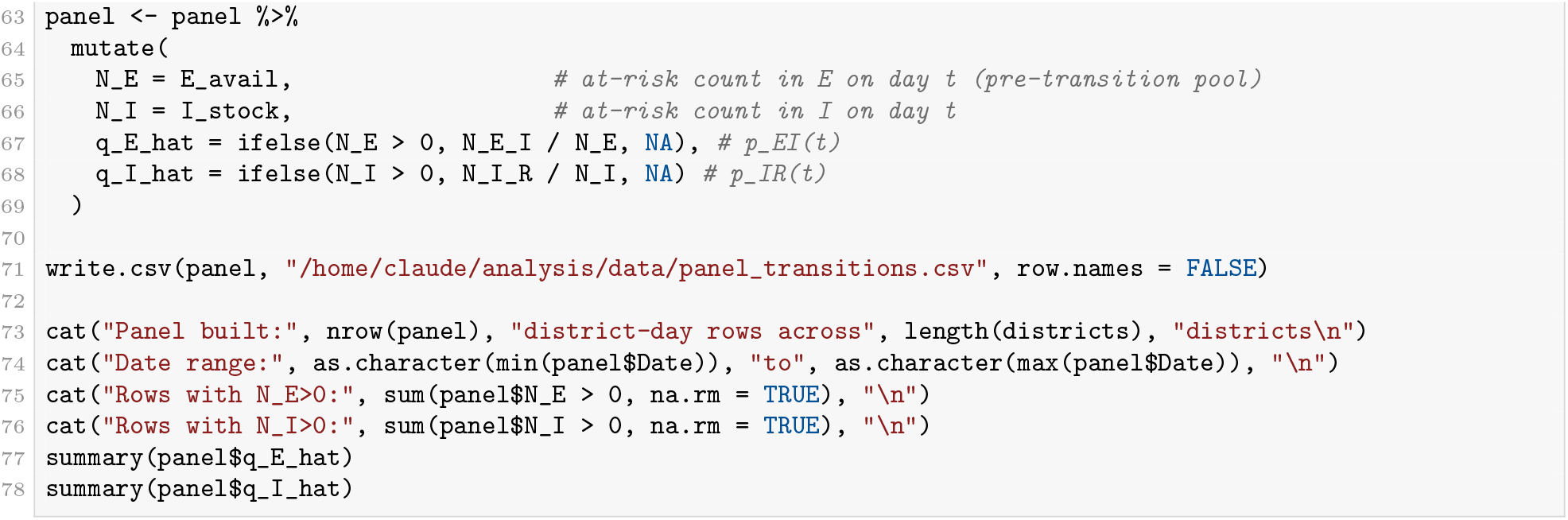

### A.2 02_modeling.R Smoothing, Confidence Intervals, LRT, GOF, and Hybrid Models

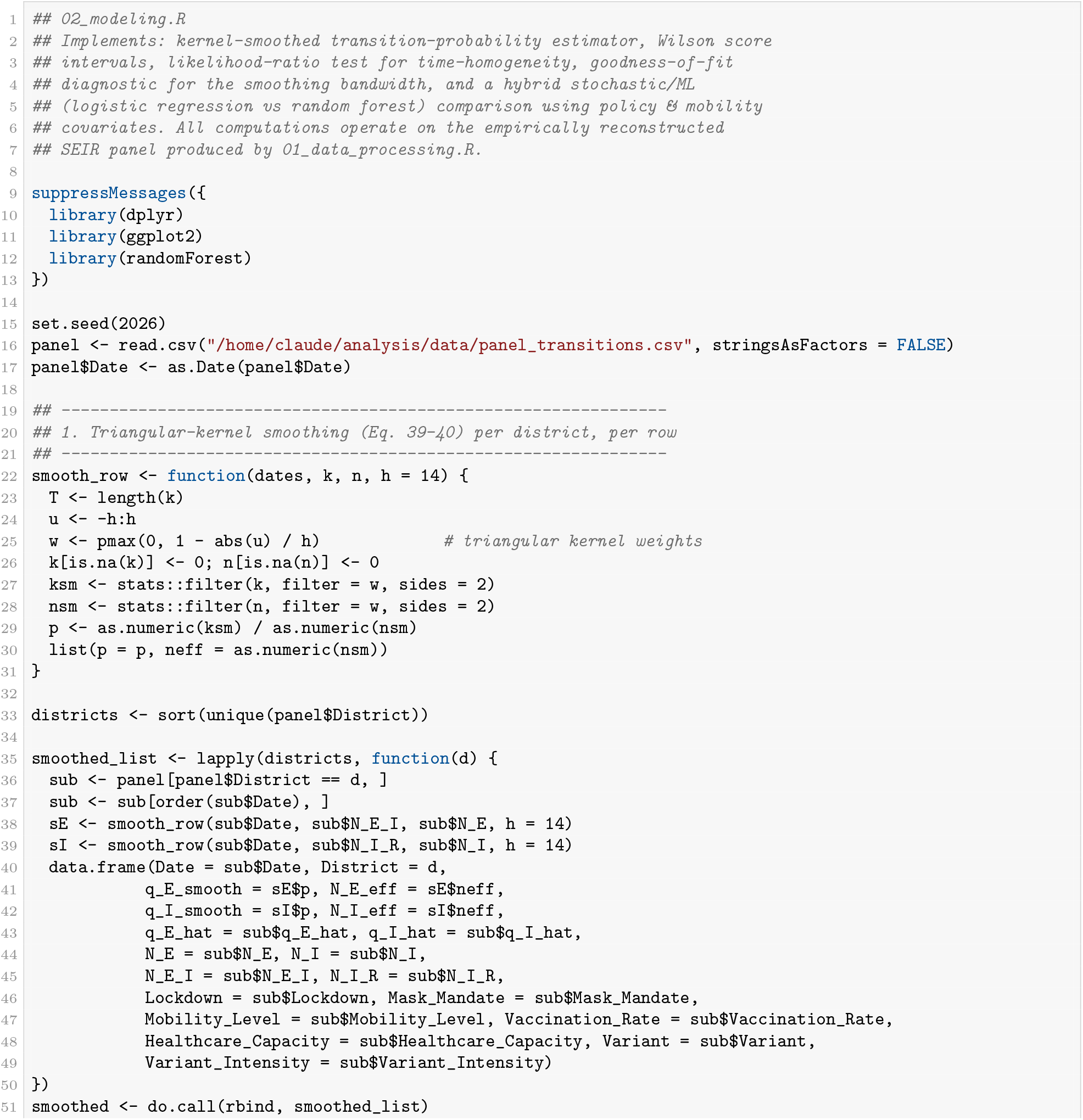
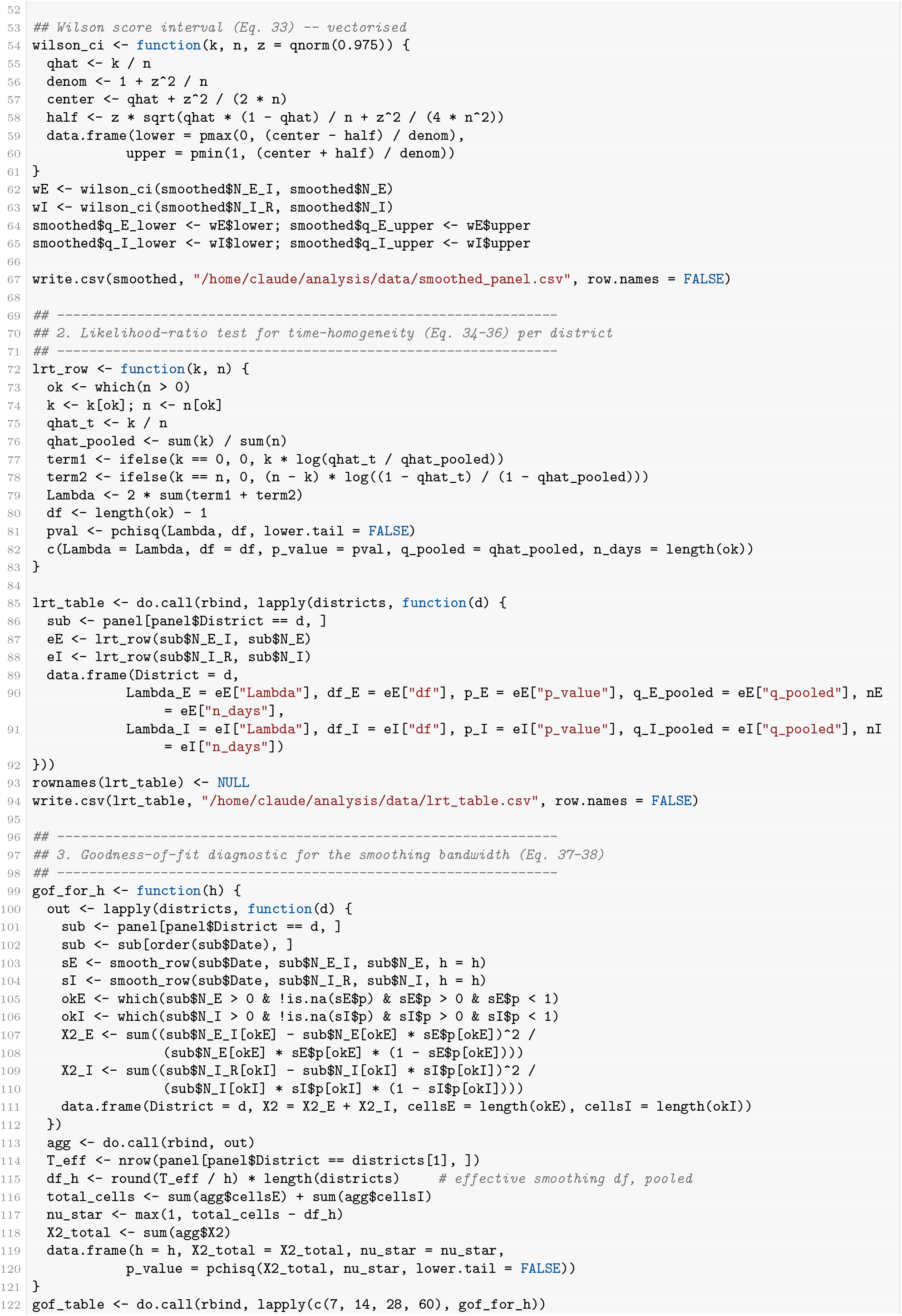
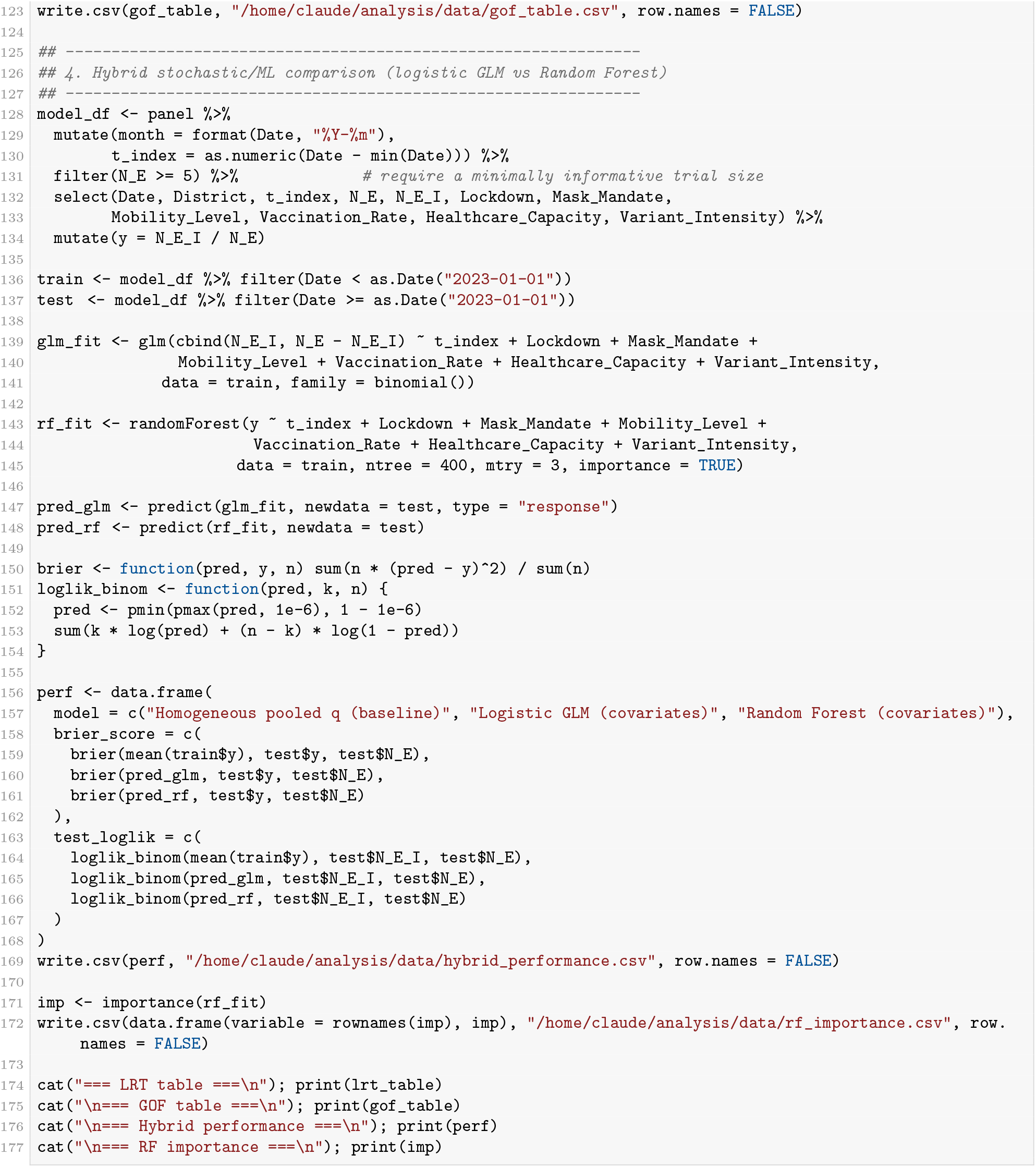

### A.3 03_figures.R Core Figures

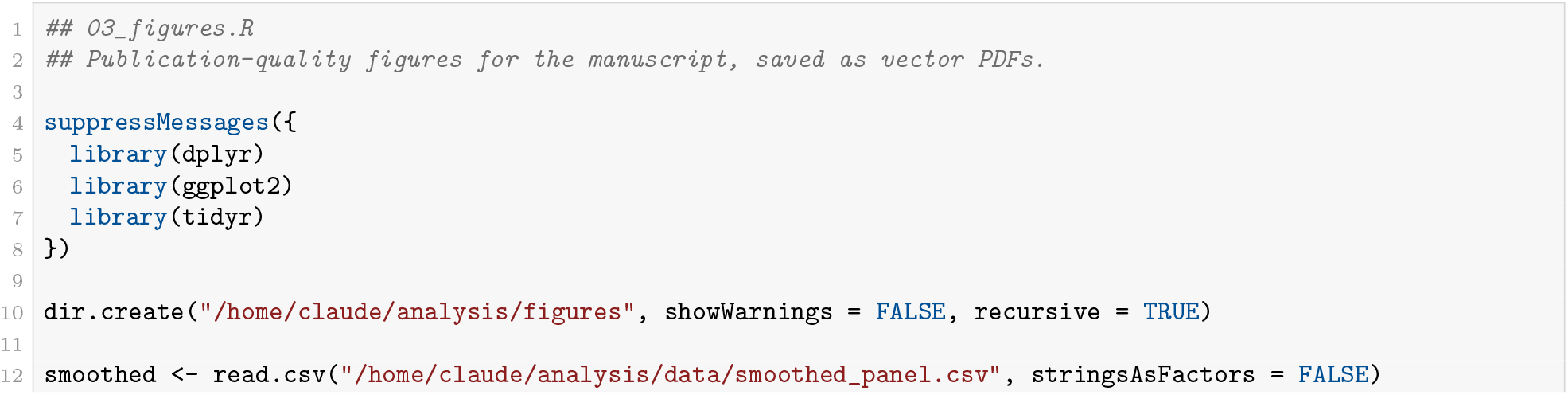
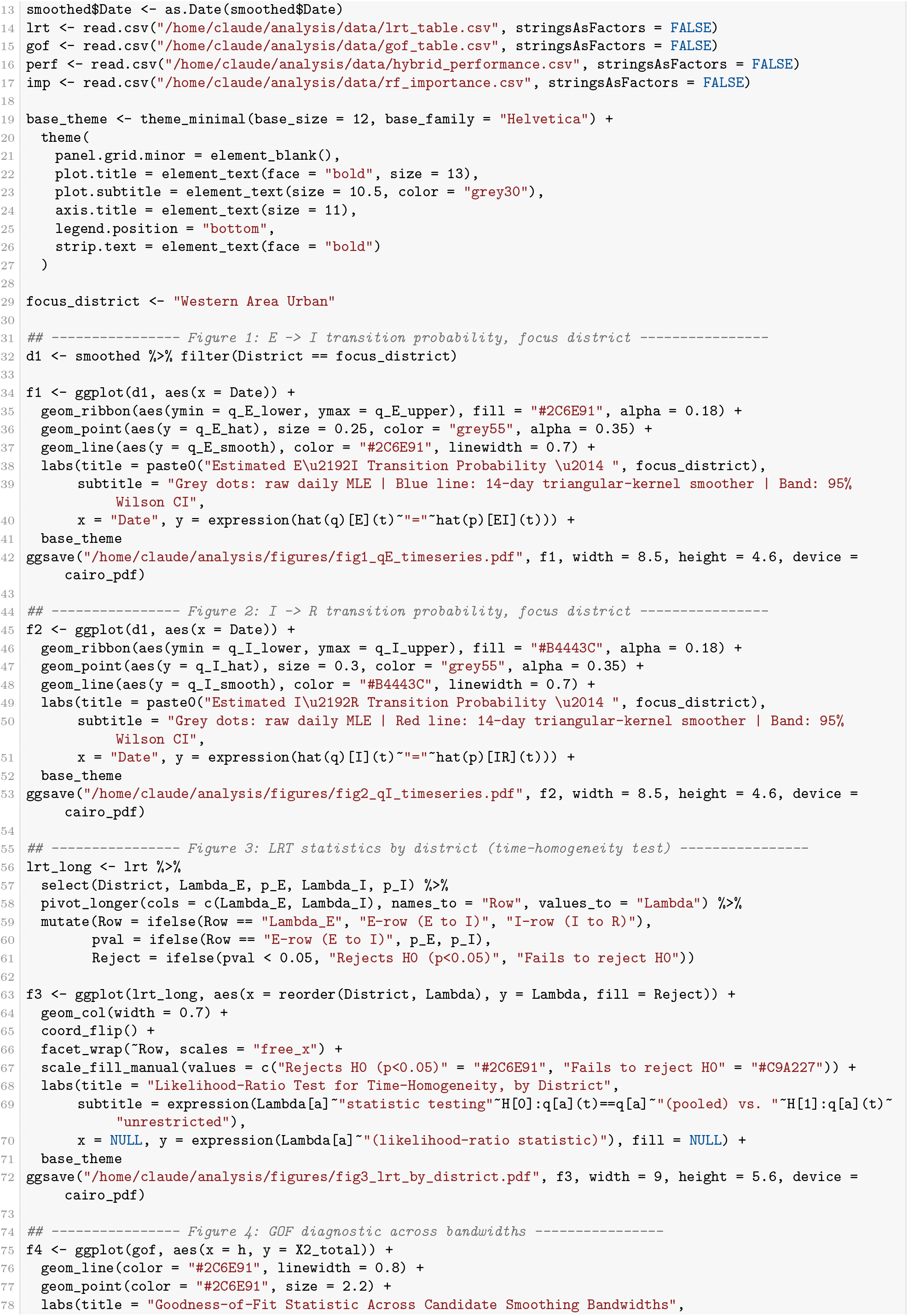
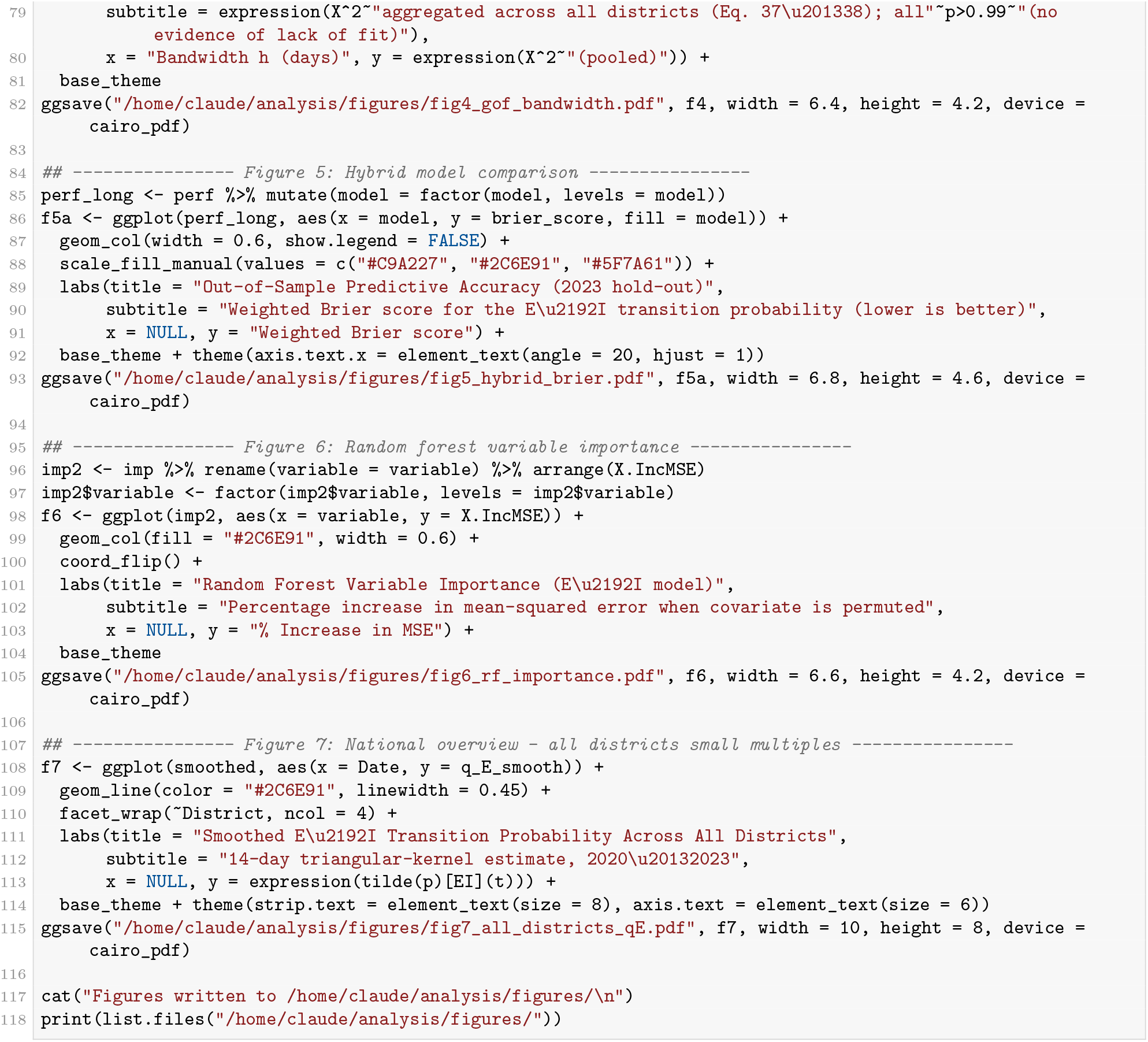

### A.4 04_full seir figures.R — Full SEIR Compartment Figures

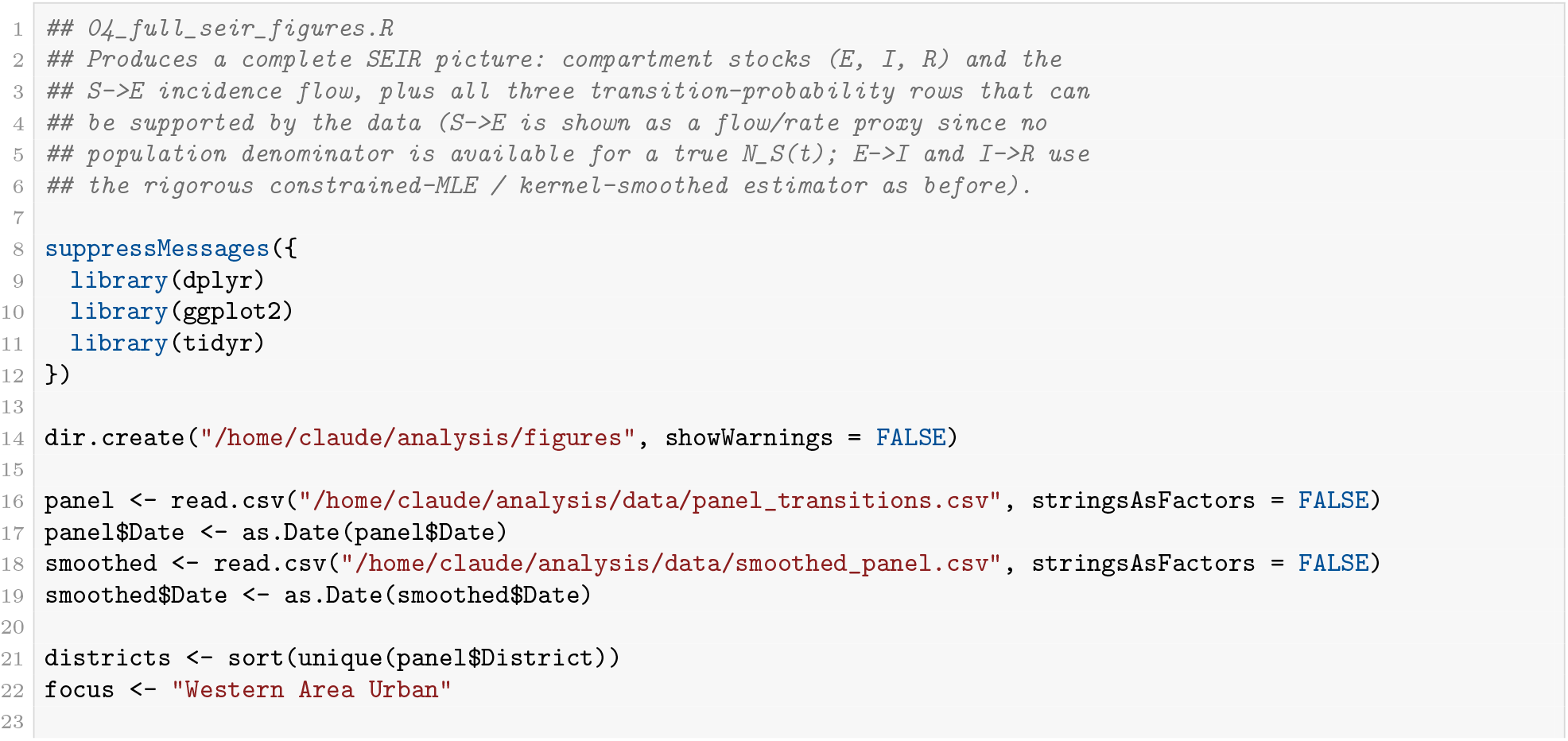
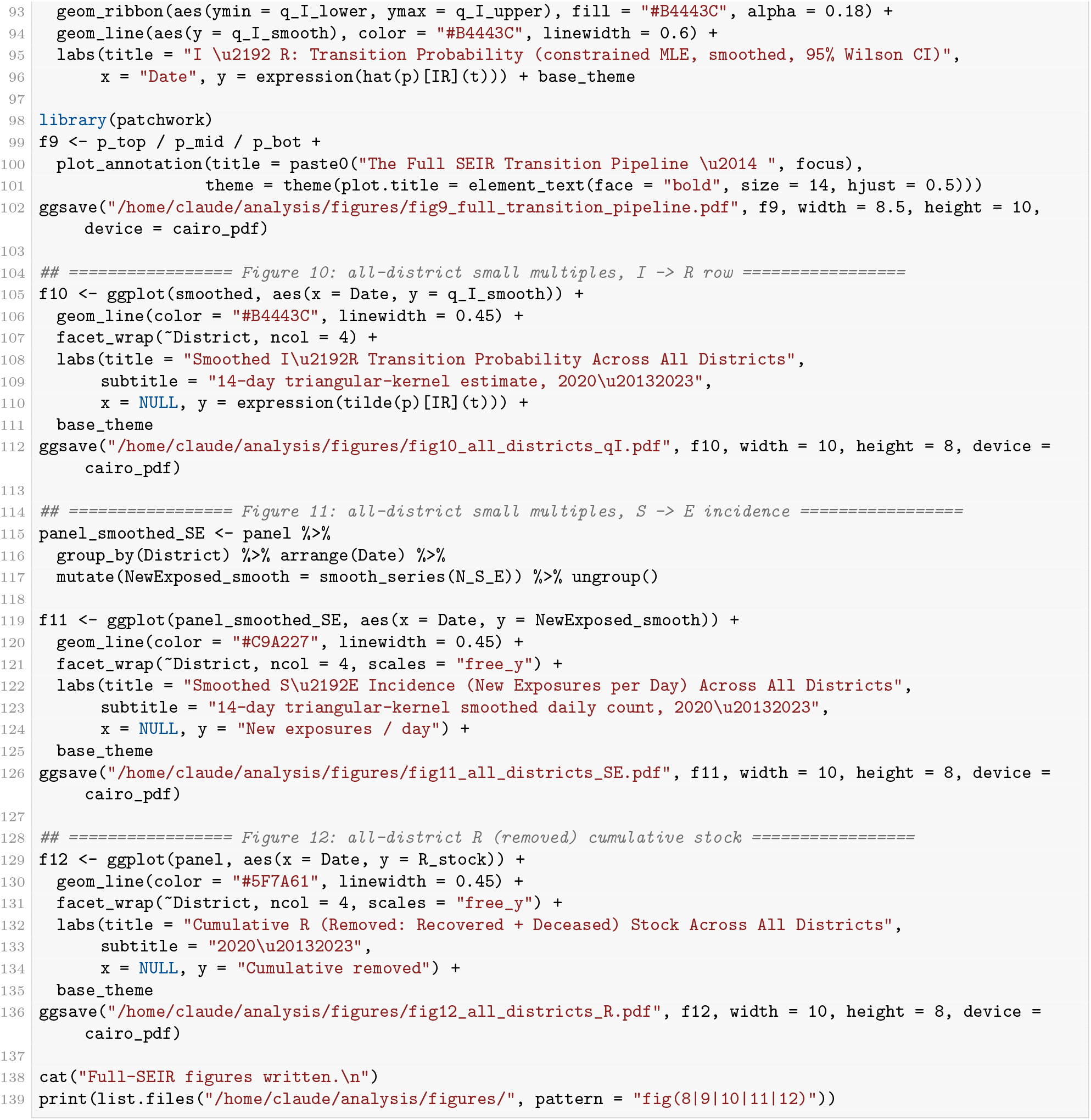
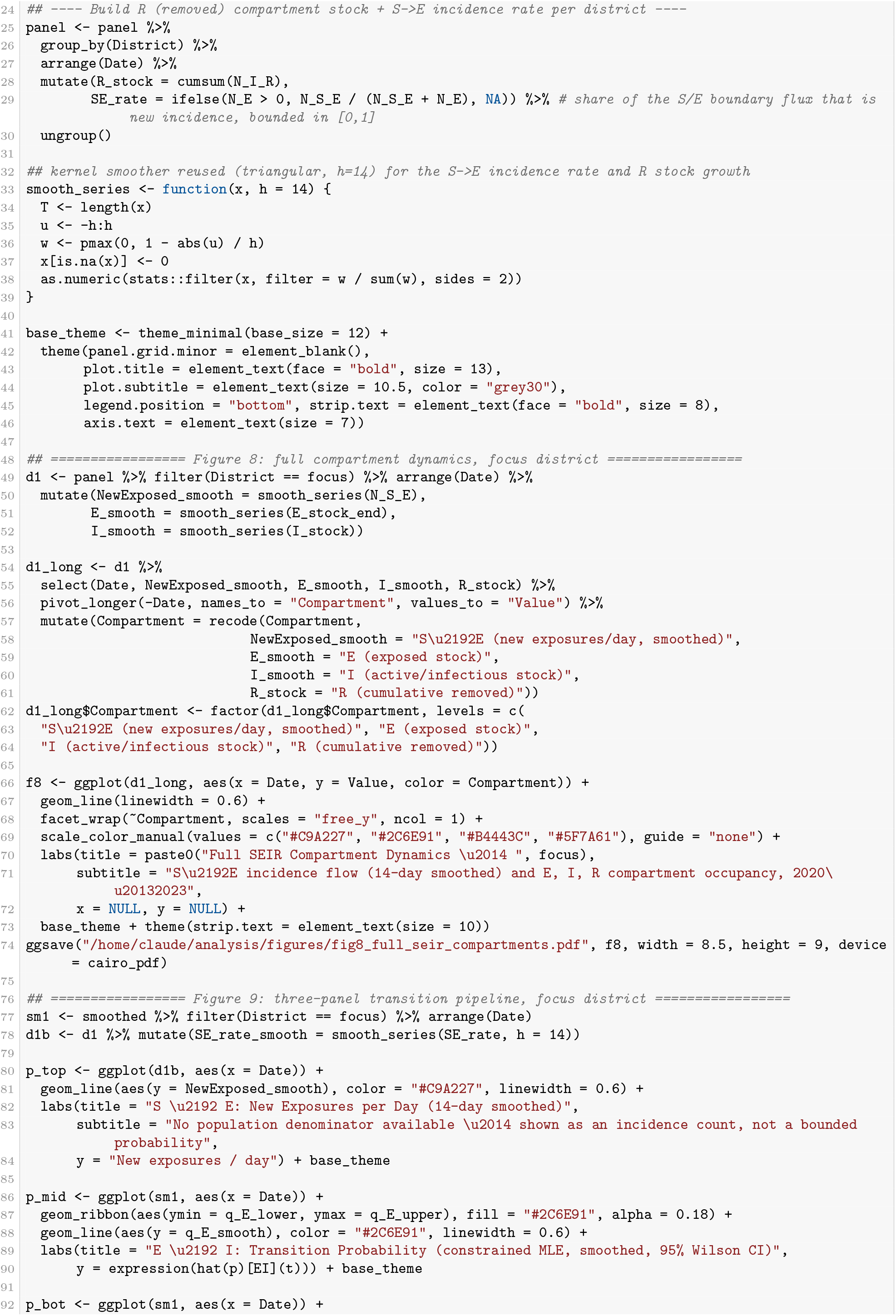

## Declaration of Generative AI and AI-Assisted Technologies

During the preparation of this work, the author(s) used ChatGPT / Claude / Gemini for language refinement, grammar editing, LaTeX code structuring, and debugging computational scripts (R/Python) for model estimation. The author(s) reviewed and edited the output as needed and take full responsibility for the content of the published article.

